# Predictive values, uncertainty, and interpretation of serology tests for the novel coronavirus

**DOI:** 10.1101/2020.06.04.20122358

**Authors:** Naomi C Brownstein, Yian Ann Chen

**Affiliations:** Moffitt Cancer Center, Department of Biostatistics and Bioinformatics, Tampa, FL, USA

## Abstract

Antibodies testing in the coronavirus era is frequently promoted, but the underlying statistics behind their validation has come under more scrutiny in recent weeks. We provide calculations, interpretations, and plots of positive and negative predictive values under a variety of scenarios. Prevalence, sensitivity, and specificity are estimated within ranges of values from researchers and antibodies manufacturers. Illustrative examples are highlighted, and interactive plots are provided in the Supplementary Material. Implications are discussed for society overall and across diverse locations with different levels of disease burden. Specifically, the proportion of positive serology tests that are false can differ drastically from up to 3% to 88% for people from different places with different proportions of infected people in the populations while the false negative rate is typically under 10%.

## 1 Introduction

The SARS-CoV-2 pandemic^1^ is wreaking havoc on physical^2^,^3^, mental^4^, economic^5–7^, and general societal health^8^,^9^. Potential treatments for Covid19 currently have limited evidence of efficacy^1^. Thus, it is critical to develop agents to prevent the spread of coronavirus, such as vaccines. Although research and development on vaccine candidates is ongoing^10^,^11^, widespread availability of a safe and effective vaccine is not expected for months or even years^12–14^. Simultaneously, there is increasing evidence of asymptomatic infection and spread^15^. With scarce testing supplies^16^, many people, blind to their prior infection status or lack thereof, are self-isolating; the current situation has even consequently been derided as Schroedinger’s virus^17^.

Without a vaccine, excitement about antibodies testing is growing.^18^ Theoretical benefits of identifying individuals with antibodies abound. Daily activities such as shopping, traveling, and dining could begin to resume, alleviating currently acute social and economic effects of the pandemic. Hoping to implement these potential benefits, some politicians have considered issuing immunity passports for people who are cleared by an antibodies test^19^. Yet, serology testing is not a panacea, and is associated with concerns about its use^20–23^. Proposals for implementing serology testing programs and understanding their benefits and limitations are available^24^,^25^

Given the increase in testing, proper interpretation of the results is critical with implications for medicine, public policy, and personal action. The goal of this paper is to estimate, quantify and visualize uncertainty in the predictive values and false positive rates of serology testing candidates available at the time of writing. Graphical displays of predictive values feature a range of scenarios. Section ^2^ reviews key metrics for serology tests. Section ^3^ summarizes and visualizes metrics in general and for serology tests operating under an Emergency Use Authorization (EUA). Section ^4^ applies the concepts to specific locations in the United States (US). Section ^5^ discusses limitations and implications. Finally, Section ^6^ details our statistical and graphical methods.

## 2 Background

This section outlines key statistical definitions related to serology testing. Readers familiar with testing characteristics may skip to Section ^3^. Definitions are included in the Supplementary Material.Additional details on these concepts and examples relevant to serology may be found elsewhere^25–28^.

Two properties of serology tests quantify how well the tests perform in on samples in a lab with known antibody status, Sensitivity is the probability that a serology test correctly classifies a sample with antibodies for SARS-CoV-2. According to the FDA, sensitivity of a test refers to its “ability to identify those with antibodies to SARS-CoV-2” and “can be estimated by determining whether or not it is able to detect antibodies in blood samples from patients who have been confirmed to have COVID-19 with a nucleic acid amplification test.” Specificity, is the probability that a test correctly classifies uninfected samples as lacking antibodies for SARS-CoV-2. Similarly, the specificity of a test refers to its “ability to identify those without antibodies to SARS-CoV-2” and is “estimated by testing large numbers of samples collected and frozen before SARS-CoV-2 is known to have circulated”. Sensitivity and specificity are pretest quantities, or validation metrics primarily meaningful before a serology test is taken. Sensitivity and specificity are defined by equations (3) and (4) in the Supplementary material. In this article, we will use these definitions unless specified otherwise.

Rather, people who confront serology tests are likely interested in post-test probabilities, including positive predictive value (PPV) and negative predictive value (NPV). PPV is the probability that a person with a positive serology test indeed has a prior infection with and antibodies for SARS-CoV-2. NPV is the probability that a person who tests negative lacks antibodies for and has not yet been infected with SARS-CoV-2. Definitions and calculations for NPV and PPV are in the Supplementary Material. PPV and NPV are more relevant to patients and clinicians in interpreting serology test results than sensitivity and specificity.

The complements of the predictive values are probabilities that test results of each type are false. The False Positive Rate (FPR), the complement of PPV, is the proportion of people who test positive that are actually lacking a prior coronavirus infection. Similarly, the false negative rate (FNR), the complement of NPV, is the proportion of people who test negative that actually had a prior infection with coronavirus. The FPR (FNR) can be interpreted as the proportion of positive (negative) serology tests are false positives (negatives). Equations for FPR and FNR are provided in the supplementary material.

## 3 Results

This section reports values for the statistics described in section 2 to help contextualize serology test results. Prevalence estimates are reported in Section 3.1. Graphical displays of NPV and PPV for the range of values under study are found in Section 3.2. Sensitivity, specificity, PPV, and FPR for tests under study are reported in 3.3.

### 3.1 Prevalence Estimates

Due to the lack of available diagnostic tests in the US, official counts of Covid-19 cases are likely undercounted^29^,^30^. Additionally, people with asymptomatic infections are unlikely to seek medical care or diagnostic testing and are likely excluded from official counts. Consequently, reliable prevalence estimates are limited. Prevalence, which affects predictive value estimates, can be considered unknown, and varies over time.

Emerging research is beginning to estimate population prevalence. Over the period from March 31 to April 7, by one estimate^31^, the prevalence by state ranged from about 0.4% in Alaska, Hawaii, Kentucky, and West Virgina to 8.5% in New York with a median prevalence of 0.9%. Another estimate^30^ from April 11 found infection proportions within states spanning from 0.1% in rural states to 7.0% in New York, and an overall US prevalence of 1.2%. A third group proposes a method^29^ with estimates that could indicate a prevalence of up to 10% of the population as of April 4, 2020. A collection of case studies is highlighted in Section ^4^. In brief, the prevalence of Covid-19 in specific cities, states, and counties estimates during the early stage of the pandemic in the spring of 2020 ranged from less than 1% to about 30% in especially hard hit areas, such as Boston and New York City. According to more recent estimates as of September 2020 by the US Centers for Disease Control and Prevention^32^, most states had seroprevalence estimates ranging from 1% to 10%. As of the writing of this paper, only four states have estimates below 1% and five have estimates exceeding 10%.

### 3.2 General Interpretation in the Context of Antibodies Testing

In this section we investigate ranges of plausible values of sensitivity, specificity for antibodies tests available and and prevalence for relevant areas and compare the predictive values. Predictive values are of interest based on minima of 80% sensitivity and 90% specificity to reflect reported values for FDA-authorized serology tests and described in Section 3.3. In general, the false negative rate is low and false positive rate is highly variable for available serology tests.

Figure 1 is a plot of NPV for these specificity, and sensitivity values and prevalence ranging from 1% to 30%, the range currently reported elsewhere as discussed in Sections 3.1 and 4. Under these scenarios, the minimum NPV was 91.3%, indicating that the false negative rate was less than 10% in all scenarios. Thus, NPV should be high, and FNR should be low for all serology tests within similar ranges for sensitivity, specificity, and prevalence. In other words, negative serology tests have a high likelihood of accurately reflecting a lack of antibodies in the general population of non-infected individuals.

**Figure 1.**
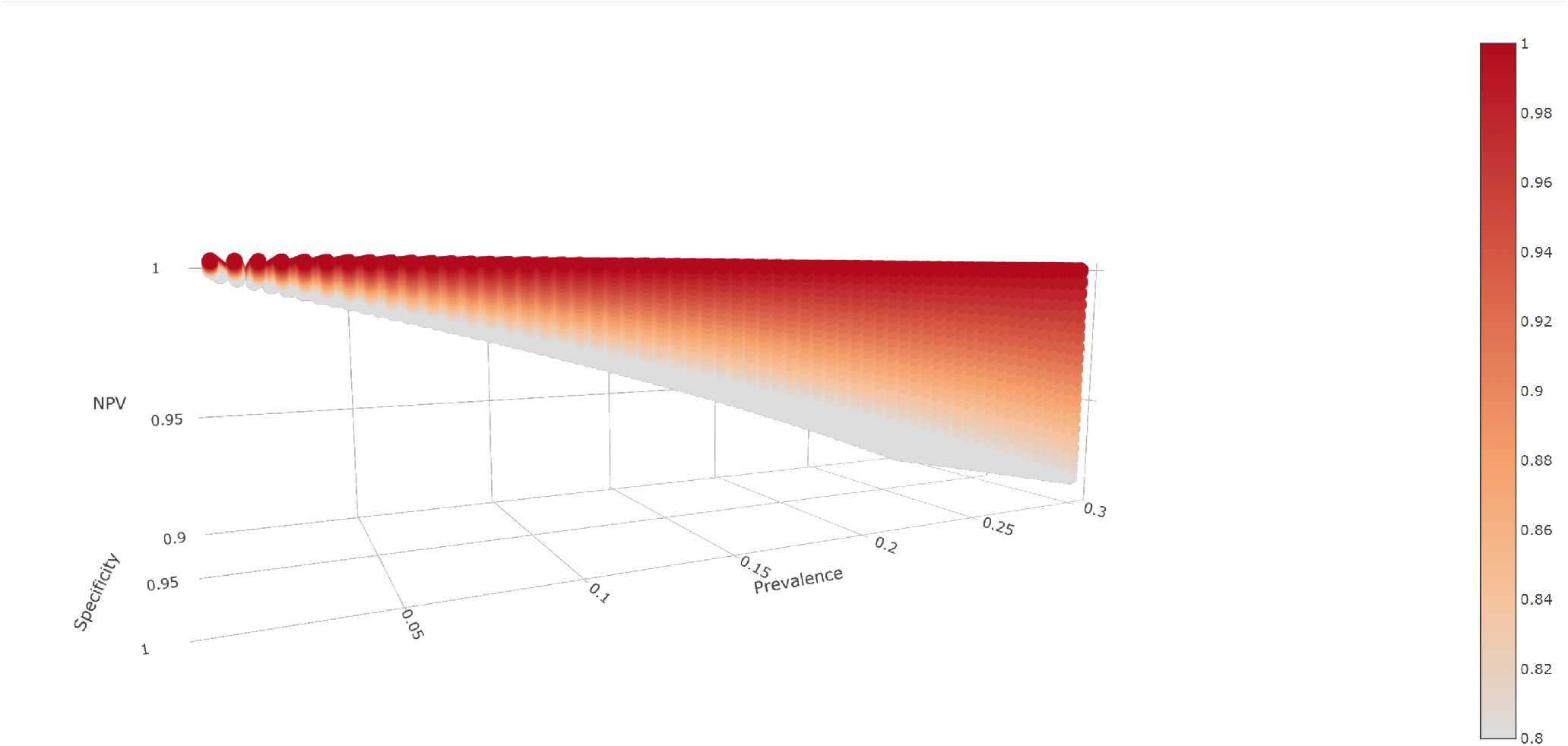
Plot of NPV by prevalence (0 to 0.3), specificity (0.9 to 1), and sensitivity (0.8 to 1). The bar on the right with sensitivity is denoted by color, with lighter colors denoting lower sensitivity and darker colors denoting higher sensitivity. All parameters are reported as decimals ranging from 0 to 1. NPV values exceeded 0.9 for all input parameters.

By contrast, figure 2 shows a corresponding plot spanning a wide range of plausible PPV values. PPV increases with prevalence and is low with lower rates of antibodies in the population. At fixed prevalence values, specificity also quickly increases PPV with prevalence. Higher sensitivity improves PPV, although the growth of PPV with increases in sensitivity is less pronounced than with increases in specificity at a given prevalence. For example, an area with 10% prevalence would have 47.1% PPV for a test with 80% sensitivity and 90% specificity, 66.7% PPV for a test with 90% sensitivity and 95% specificity, and 91.3% PPV for a test with 95% sensitivity and 99% specificity. Equivalently, the false positive rates would be 52.9%, 33.3%, and 8.7%. In areas with a 30% infection rate, the same tests would yield respective PPVs of 77.4%, 88.5% and 97.6%, and false positives of 22.6%, 11.5%, and 2.4%. Yet, if the prevalence is 1%, then PPV could reach 49.0% for 95% sensitivity and 99% specificity or fall as low as 7.5%, indicating that only 7.5% of people with positive serology tests in fact possess antibodies. Put another way, for tests with 80% sensitivity and 90% specificity in locations with 1% prevalence, about 93% of people with positive serology tests would be expected to <u>lack</u> antibodies for SARS-CoV-2!

**Figure 2.**
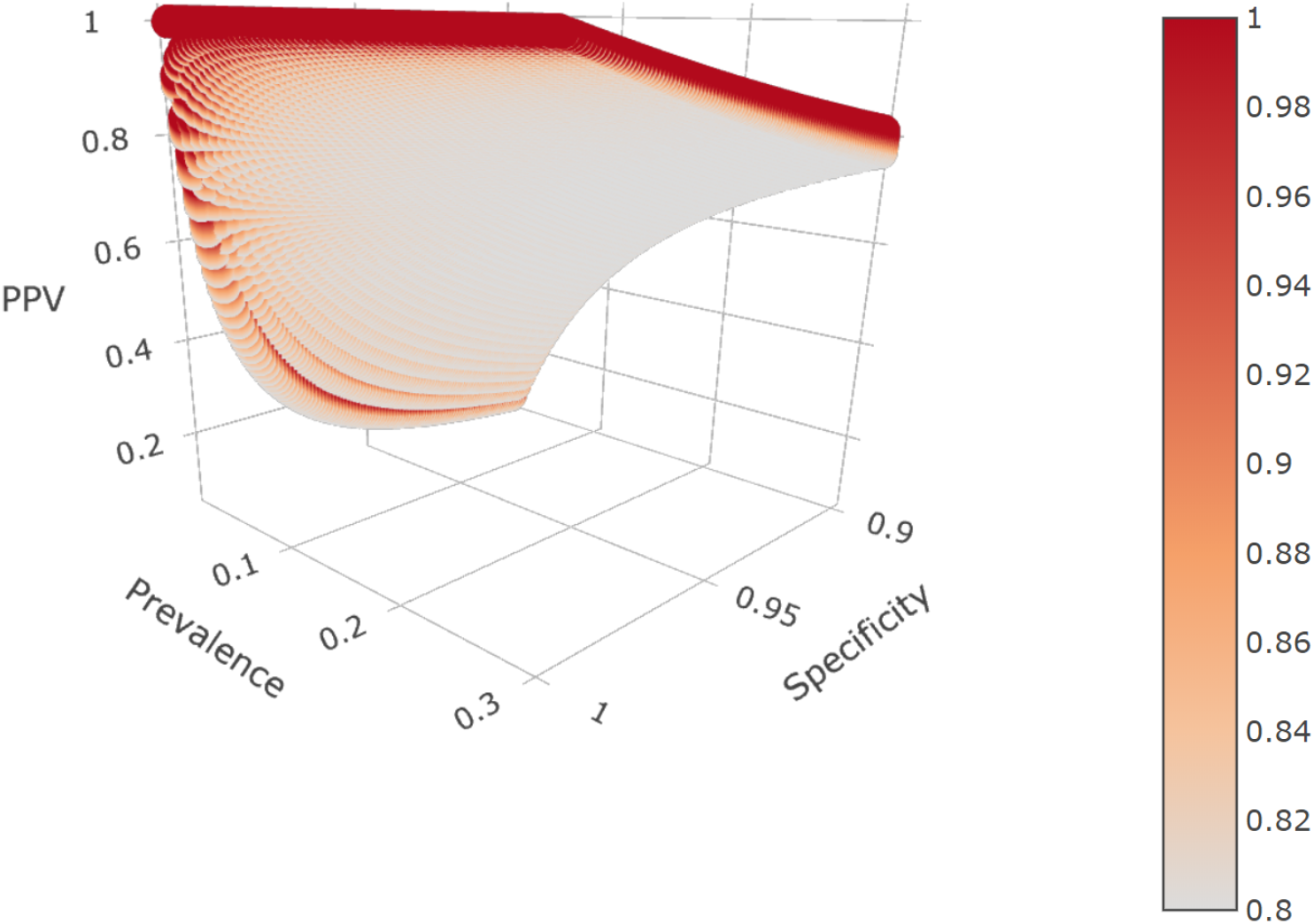
Plot of PPV by prevalence (0.01 to 0.3), specificity (0.9 to 1), and sensitivity (0.8 to 1). Sensitivity is denoted by color, with lighter colors denoting low sensitivity and darker colors denoting higher sensitivity. All parameters are reported as decimals ranging from 0 to 1. PPV varied widely based on different parameters, ranging from 0.07 to 1.

### 3.3 Analysis of Individual Antibodies Tests

As of May 22, 2020, the FDA had allowed 13 serology tests to operate under an Emergency Use Authorization (EUA)^33^. By November 24, 2020, the total number of tests with EUA had increased^34^ to 89, and EUA had been revoked for two prior tests. However, some of the updated results are repetitive, with many companies reporting test characteristics for IgG, IgM, and Combined(IgG/IgM). In these cases, to balance space and conciseness with completeness, we reported the combined tests only. Characteristics of these 61 distinct tests appear in Table 1. Sensitivity estimates range from about 88% to 100%, with 95% confidence limits ranging from about 74% to 100%. Specificity estimates range from about 95% to 100%, with confidence bounds ranging from 88% to 100%.

**Table 1.** Statistics for FDA authorized serology tests with EUA All numbers are percentages. Interval estimates for sensitivity and specificity are 95% confidence intervals reported by the FDA^33^. Interval estimates for the false positive rates are minimum and maximum values of all possible false positive rates calculated at the specified prevalence level for the corresponding test based on all possible estimates of sensitivity and specificity within the 95% confidence intervals. \**n* refers to the number of samples used to calculate the pretest probabilities. The number on the left refers to the number of samples with infected with SARS-COV-2 that were tested to estimate sensitivity. The number on the right refers to the number of control samples used to estimate specificity. Test types are shortened as followed. “G” denotes IgG, “M” denotes IgM, “C” denotes combined IgG/IgM, and “P” denotes Pan-IgG.

| Test | Type | Sensitivity | Specificity | n (sen, spec) | FPR 1% | FPR 5% | FPR 10% |
| --- | --- | --- | --- | --- | --- | --- | --- |
| Abbott-AI | G | 100.0 (89.9, 100.0) | 99.0 (94.6, 99.8) | 34;100 | 49.7 (16.5, 85.6) | 16.0 (3.7, 53.3) | 8.3 (1.8, 35.1) |
| Abbott-Ar | G | 100.0 (95.8, 100.0) | 99.6 (99.0, 99.9) | 88;1070 | 28.4 (9.0, 50.8) | 7.1 (1.9, 16.6) | 3.5 (0.9, 8.6) |
| Abbott-AdvDxAI | M | 95.0 (83.5, 98.6) | 99.6 (99.3, 99.7) | 40;2985 | 29.4 (23.1, 45.4) | 7.4 (5.5, 13.7) | 3.7 (2.7, 7.0) |
| Abbott-AdviseDxAr | M | 95.0 (83.5, 98.6) | 99.6 (99.3, 99.7) | 40;2965 | 29.4 (23.1, 45.4) | 7.4 (5.5, 13.7) | 3.7 (2.7, 7.0) |
| AccessBio | C | 98.4 (91.7, 99.7) | 98.9 (96.1, 99.7) | 64;182 | 52.5 (23.0, 80.8) | 17.5 (5.4, 44.7) | 9.1 (2.6, 27.7) |
| AssureTech | C | 100.0 (88.7, 100.0) | 98.8 (93.3, 99.8) | 30;80 | 54.3 (16.5, 88.2) | 18.6 (3.7, 58.9) | 9.7 (1.8, 40.5) |
| Babson | G | 100.0 (88.3, 100.0) | 100.0 (96.3, 100.0) | 29;100 | 0.0 (0.0, 80.6) | 0.0 (0.0, 44.3) | 0.0 (0.0, 27.4) |
| BeckmanCoulter | G | 96.8 (91.1, 98.9) | 99.6 (99.2, 99.8) | 95;1400 | 29.0 (16.7, 46.5) | 7.3 (3.7, 14.3) | 3.6 (1.8, 7.3) |
| BeckmanCoulter | M | 96.7 (92.5, 98.6) | 99.9 (99.5, 100.0) | 151;1400 | 9.3 (0.0, 34.9) | 1.9 (0.0, 9.3) | 0.9 (0.0, 4.6) |
| Beijing W ELISA | P | 96.7 (83.3, 99.4) | 97.5 (91.3, 99.3) | 30;80 | 71.9 (41.1, 91.2) | 32.9 (11.8, 66.5) | 18.9 (6.0, 48.5) |
| Beijing W Rapid | P | 100.0 (88.7, 100.0) | 98.8 (93.3, 99.8) | 30;80 | 54.3 (16.5, 88.2) | 18.6 (3.7, 58.9) | 9.7 (1.8, 40.5) |
| Bio-Rad | P | 98.0 (89.5, 99.6) | 99.3 (98.3, 99.7) | 50;600 | 41.4 (23.0, 65.3) | 11.9 (5.4, 26.5) | 6.0 (2.6, 14.6) |
| BioCan | C | 93.3 (78.7, 98.2) | 96.2 (89.4, 98.7) | 30;79 | 80.1 (56.7, 93.0) | 43.6 (20.1, 71.9) | 26.8 (10.6, 54.8) |
| BioCheck | C | 99.1 (95.0, 99.8) | 97.2 (93.0, 98.9) | 110;143 | 73.7 (52.2, 87.9) | 34.9 (17.3, 58.3) | 20.3 (9.0, 39.9) |
| Biohit | C | 96.7 (83.3, 99.4) | 95.0 (87.8, 98.0) | 30;80 | 83.7 (66.6, 93.5) | 49.6 (27.7, 73.6) | 31.8 (15.3, 56.9) |
| bioMerieux | G | 100.0 (88.3, 100.0) | 99.9 (99.4, 100.0) | 29;989 | 9.0 (0.0, 40.2) | 1.9 (0.0, 11.4) | 0.9 (0.0, 5.8) |
| bioMerieux | M | 100.0 (85.7, 100.0) | 99.4 (97.7, 99.8) | 23;308 | 37.3 (16.5, 72.7) | 10.2 (3.7, 33.8) | 5.1 (1.8, 19.5) |
| Cellex | C | 93.8 (88.2, 96.8) | 96.0 (92.8, 97.8) | 128;250 | 80.8 (69.2, 89.0) | 44.8 (30.2, 60.8) | 27.7 (17.0, 42.4) |
| DiaSorin | G | 97.6 (87.4, 99.6) | 99.3 (98.6, 99.6) | 41;1090 | 41.5 (28.4, 61.3) | 12.0 (7.1, 23.3) | 6.1 (3.5, 12.6) |
| DiaSorin | M | 91.8 (85.6, 95.5) | 99.3 (98.9, 99.5) | 122;2473 | 43.0 (34.1, 56.0) | 12.7 (9.0, 19.6) | 6.4 (4.5, 10.4) |
| Diazyme | G | 100.0 (88.3, 100.0) | 97.4 (96.1, 98.3) | 29;852 | 72.0 (62.7, 81.4) | 33.1 (24.4, 45.6) | 19.0 (13.3, 28.4) |
| Diazyme | M | 94.4 (88.4, 97.4) | 98.3 (96.2, 99.3) | 108;302 | 64.1 (41.6, 81.0) | 25.5 (12.0, 45.0) | 13.9 (6.1, 27.9) |
| Emory | G | 100.0 (88.7, 100.0) | 96.4 (94.6, 97.6) | 30;638 | 78.1 (70.4, 85.8) | 40.6 (31.3, 53.6) | 24.5 (17.8, 35.4) |
| EUROIMMUN | G | 90.0 (74.4, 96.5) | 100.0 (95.4, 100.0) | 30;80 | 0.0 (0.0, 86.0) | 0.0 (0.0, 54.0) | 0.0 (0.0, 35.8) |
| Genalyte | P | 96.1 (92.2, 98.1) | 97.7 (96.4, 98.5) | 181;862 | 70.3 (60.2, 79.4) | 31.3 (22.5, 42.6) | 17.7 (12.1, 26.0) |
| GenScript | P | 100.0 (87.1, 100.0) | 100.0 (95.8, 100.0) | 26;88 | 0.0 (0.0, 82.7) | 0.0 (0.0, 47.8) | 0.0 (0.0, 30.3) |
| HangzhouRapid | C | 100.0 (88.7, 100.0) | 100.0 (95.4, 100.0) | 30;80 | 0.0 (0.0, 83.7) | 0.0 (0.0, 49.6) | 0.0 (0.0, 31.8) |
| HangzhouLaihe | C | 100.0 (88.7, 100.0) | 98.8 (93.3, 99.8) | 30;80 | 54.3 (16.5, 88.2) | 18.6 (3.7, 58.9) | 9.7 (1.8, 40.5) |
| Healgen | C | 100.0 (88.7, 100.0) | 97.5 (91.3, 99.3) | 30;80 | 71.2 (40.9, 90.7) | 32.2 (11.7, 65.1) | 18.4 (5.9, 46.9) |
| InBios | G | 100.0 (88.7, 100.0) | 100.0 (95.4, 100.0) | 30;80 | 0.0 (0.0, 83.7) | 0.0 (0.0, 49.6) | 0.0 (0.0, 31.8) |
| InBios | M | 96.7 (83.3, 99.4) | 98.8 (93.3, 99.8) | 30;80 | 55.1 (16.6, 88.8) | 19.1 (3.7, 60.4) | 10.0 (1.8, 42.0) |
| JiangSu | C | 100.0 (93.8, 100.0) | 94.8 (88.5, 97.8) | 58;97 | 83.7 (68.5, 92.4) | 49.7 (29.5, 70.0) | 31.9 (16.5, 52.5) |
| Luminex | G | 96.3 (89.8, 98.7) | 99.3 (98.3, 99.7) | 82;603 | 41.8 (23.1, 65.2) | 12.1 (5.5, 26.5) | 6.1 (2.7, 14.6) |
| Megna | C | 100.0 (88.7, 100.0) | 95.0 (87.8, 98.0) | 30;80 | 83.2 (66.4, 93.2) | 48.7 (27.5, 72.3) | 31.0 (15.3, 55.3) |
| Mt Sinai | C | 92.5 (80.1, 97.4) | 100.0 (95.1, 100.0) | 40;74 | 0.0 (0.0, 85.8) | 0.0 (0.0, 53.8) | 0.0 (0.0, 35.5) |
| NanoEnTek | C | 96.7 (83.3, 99.4) | 98.8 (93.3, 99.8) | 30;80 | 55.1 (16.6, 88.8) | 19.1 (3.7, 60.4) | 10.0 (1.8, 42.0) |
| Nirmidas | M | 93.1 (83.6, 97.3) | 97.9 (92.8, 99.4) | 58;97 | 69.1 (37.9, 89.5) | 30.0 (10.5, 62.1) | 16.9 (5.3, 43.7) |
| Nirmidas | G | 87.9 (77.1, 94.0) | 100.0 (96.2, 100.0) | 58;97 | 0.0 (0.0, 83.0) | 0.0 (0.0, 48.4) | 0.0 (0.0, 30.7) |
| Nirmidas | C | 96.6 (88.3, 99.1) | 97.9 (92.8, 99.4) | 58;97 | 68.3 (37.5, 89.0) | 29.2 (10.3, 60.8) | 16.4 (5.2, 42.3) |
| Ortho-Clinical | G | 90.0 (76.9, 96.0) | 100.0 (99.1, 100.0) | 40;407 | 0.0 (0.0, 53.7) | 0.0 (0.0, 18.2) | 0.0 (0.0, 9.5) |
| Ortho-Clinical | P | 100.0 (92.7, 100.0) | 100.0 (99.0, 100.0) | 49;400 | 0.0 (0.0, 51.6) | 0.0 (0.0, 17.0) | 0.0 (0.0, 8.8) |
| QuanSys | G | 95.5 (78.2, 99.2) | 99.7 (98.8, 99.9) | 22;585 | 23.7 (9.1, 60.3) | 5.6 (1.9, 22.6) | 2.7 (0.9, 12.1) |
| Quotient | P | 93.0 (85.6, 96.8) | 99.8 (98.6, 100.0) | 86;408 | 17.6 (0.0, 61.8) | 3.9 (0.0, 23.7) | 1.9 (0.0, 12.8) |
| Roche | P | 100.0 (88.3, 100.0) | 99.8 (99.7, 99.9) | 29;5272 | 16.5 (9.0, 25.2) | 3.7 (1.9, 6.1) | 1.8 (0.9, 3.0) |
| Salofa | C | 93.3 (78.7, 98.2) | 98.8 (93.3, 99.8) | 30;80 | 56.0 (16.8, 89.4) | 19.6 (3.7, 61.8) | 10.4 (1.8, 43.4) |
| Shenzhen | C | 100.0 (97.4, 100.0) | 98.7 (96.2, 99.5) | 142;226 | 56.3 (33.1, 79.4) | 19.8 (8.7, 42.6) | 10.5 (4.3, 26.0) |
| SiemensADVIA | G | 100.0 (91.6, 100.0) | 99.9 (99.6, 99.9) | 42;1831 | 9.0 (9.0, 30.2) | 1.9 (1.9, 7.7) | 0.9 (0.9, 3.8) |
| SiemensADVIA | P | 100.0 (92.4, 100.0) | 99.8 (99.4, 99.9) | 47;1589 | 16.5 (9.0, 39.1) | 3.7 (1.9, 11.0) | 1.8 (0.9, 5.5) |
| SiemensAtellica | G | 100.0 (91.6, 100.0) | 99.9 (99.7, 100.0) | 42;1841 | 9.0 (0.0, 24.5) | 1.9 (0.0, 5.9) | 0.9 (0.0, 2.9) |
| SiemensAtellica | P | 100.0 (91.6, 100.0) | 99.8 (99.3, 99.9) | 42;1091 | 16.5 (9.0, 43.1) | 3.7 (1.9, 12.7) | 1.8 (0.9, 6.4) |
| SiemensDimEXL | P | 100.0 (95.4, 100.0) | 99.9 (99.5, 100.0) | 79;1529 | 9.0 (0.0, 34.2) | 1.9 (0.0, 9.1) | 0.9 (0.0, 4.5) |
| SiemensDimVista | P | 100.0 (95.4, 100.0) | 99.8 (99.4, 99.9) | 79;1529 | 16.5 (9.0, 38.4) | 3.7 (1.9, 10.7) | 1.8 (0.9, 5.4) |
| Sugentech | G | 96.7 (83.3, 99.4) | 100.0 (95.4, 100.0) | 30;80 | 0.0 (0.0, 84.5) | 0.0 (0.0, 51.2) | 0.0 (0.0, 33.2) |
| TBG | C | 93.3 (78.7, 98.2) | 95.0 (87.8, 98.0) | 30;80 | 84.1 (66.8, 93.9) | 50.5 (27.9, 74.7) | 32.5 (15.5, 58.2) |
| ThermoFisher | P | 96.7 (83.3, 99.4) | 97.5 (91.3, 99.3) | 30;80 | 71.9 (41.1, 91.2) | 32.9 (11.8, 66.5) | 18.9 (6.0, 48.5) |
| UnivAZ | P | 97.5 (87.1, 99.6) | 99.1 (97.3, 99.7) | 40;320 | 47.7 (23.0, 75.4) | 14.9 (5.4, 37.1) | 7.7 (2.6, 21.8) |
| Vibrant | C | 98.1 (90.1, 99.7) | 98.6 (97.1, 99.3) | 53;501 | 58.6 (41.0, 76.1) | 21.3 (11.8, 37.9) | 11.4 (5.9, 22.5) |
| Wadsworth | P | 88.0 (80.5, 92.8) | 98.8 (97.3, 99.5) | 108;433 | 57.4 (34.8, 76.9) | 20.6 (9.3, 38.9) | 10.9 (4.6, 23.2) |
| Xiamen | C | 100.0 (88.7, 100.0) | 96.2 (89.5, 98.7) | 30;80 | 79.0 (56.3, 92.1) | 41.9 (19.8, 69.2) | 25.5 (10.5, 51.6) |
| Zeus | G | 93.3 (78.7, 98.2) | 100.0 (94.8, 100.0) | 30;70 | 0.0 (0.0, 86.7) | 0.0 (0.0, 55.7) | 0.0 (0.0, 37.3) |

Figures 3 and 4 highlight the relationship between PPV and prevalence for each of the 11 serology tests granted an EUA by the US FDA in the spring of 2020 that retained approval in November 2020. Similar figures for the remaining 50 tests are provided in the Supplementary material. The figures illustrate the known relationship that PPV should be lower in populations with lower prevalence^28^, and that PPV increases more rapidly with increasing specificity than with increasing sensitivity.

**Figure 3.**
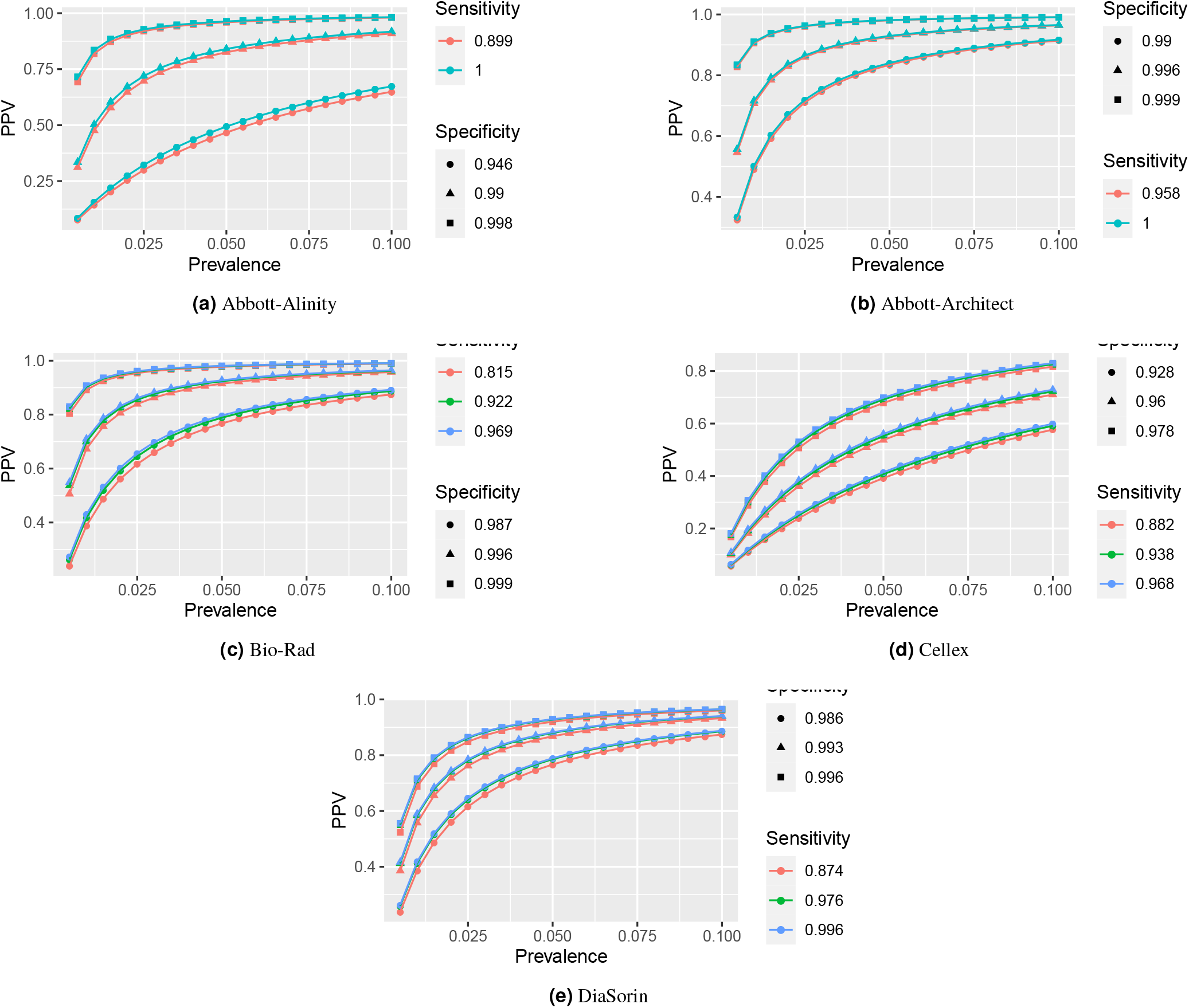
PPV by prevalence (up to 10%) for FDA tests (A-D)

**Figure 4.**
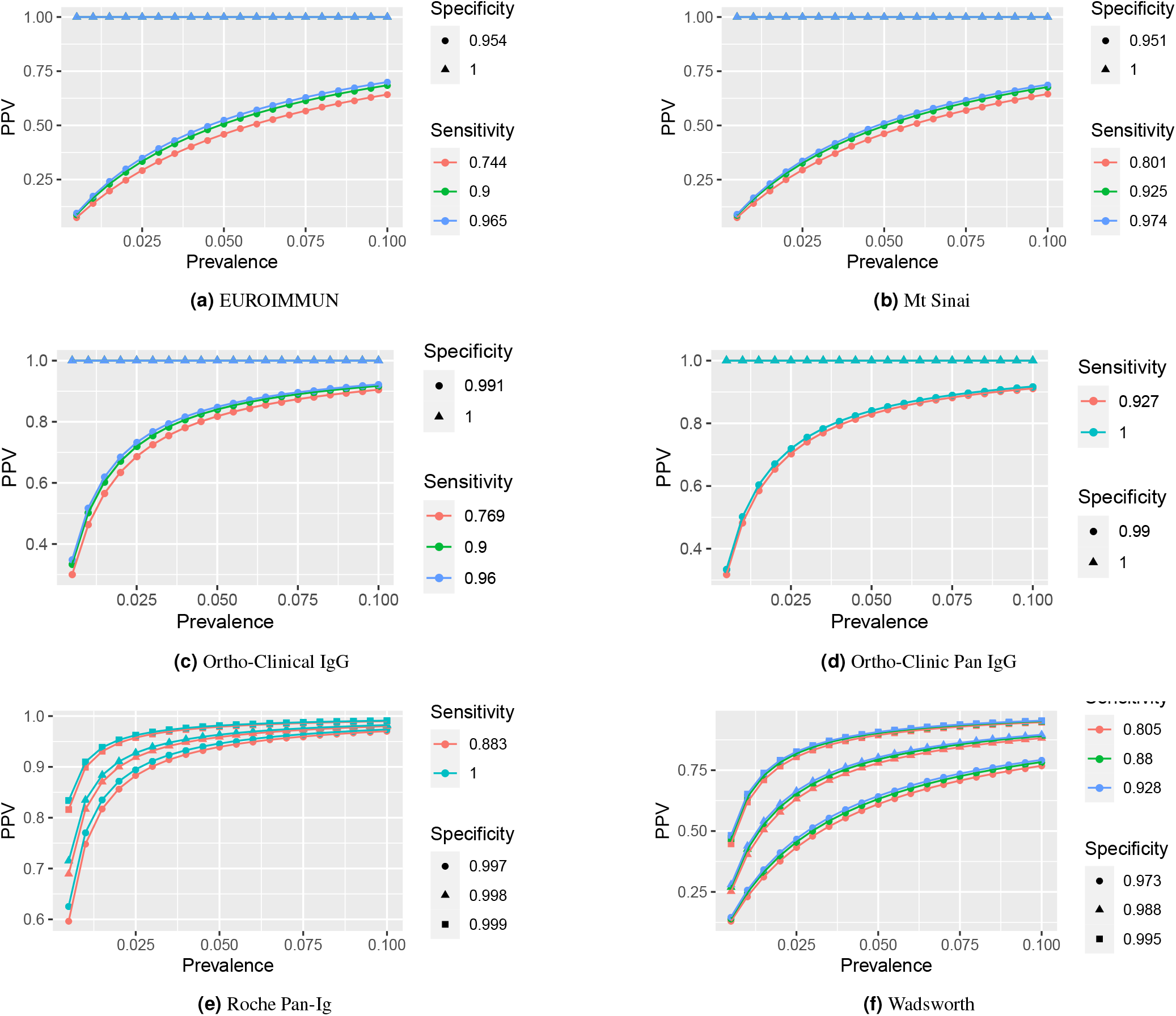
PPV by prevalence (up to 10%) for FDA tests (E-Z)

At low prevalence, such as 1%, many tests exhibit low PPV and high rates of false positives (Table 1). The upper limits of false positive rates for 45 (74%) tests exceed 60% and 49 (80%) tests exceed 50% given 1% prevalence. Thus, in regions with 1% prevalence, scenarios having 50% or more percent of positive serology tests corresponding to people *lacking* SARS-Cov-2 antibodies would be within the realm of reasonable expectations. In addition, some tests with estimated specificity below 97% have not only poor estimated false positive rates, but report high precision. Tests by Cellex, Megna, Biohit, JiangSu, and TBG are estimated to have about 80% false positives at 1% prevalence, with even the lower bounds on their FPR exceeding 66%. In such low prevalence populations, on average, anywhere between 6 and 9 out of 10 positive tests by these companies are expected to be false.

Results for all tests improve with prevalence, but overall false positive rates remain concerning. Although only nine point estimates for false positive rates exceed 20% assuming 10% prevalence, which is higher than most currently estimated infection rates^30–32^, the upper bound for false positive rates exceed 20% for 39 out of 61 tests. Moreover, only 15 (25%) tests from six companies – Abbott (3 of 4 minus Alinity IgG, Ortho-clinical (IgG and PanIg), Siemens (all 6 varieties), BeckmanCoulter (IgG and IgM), BioMerieux (IgG) and Roche – have upper bounds on false positive rates below 10% when the prevalence is 10%. In other words, 46 out of 61 tests could reasonably produce at least 1 false positive out of 10 positive tests if the regional prevalence is 10%.

Estimates or confidence bounds of exactly 100%, reported for many, are shown for completeness. These estimates should be interpreted with caution, as tests establishing sensitivity and specificity were done on small samples^33^, which may lack sufficient precision^20^,^35^ for estimation when these quantities are expected to be close to 1. For example, while specificity estimates of the tests by Ortho-Clinical were validated based on samples of around 400, estimates of 100% sensitivity by Abbott and Roche or 100% specificity by Euroimmun and Mt. Sinai were validated on samples of less than 100. Yet, as shown by the definitions and equations in the Supplementary Material, perfect (100%) specificity implies perfect PPV and perfect sensitivity implies perfect NPV. In these cases, it is especially important to consider the entire range of values for PPV and FPR. Indeed, while tests by Mt. Sinai and Euroimmun have point estimates and lower interval estimates of 0% false positives, upper interval estimates climb to about 86%. Even at 10% prevalence, if the true specificity is closer to the lower 95% confidence limit, then false positive rate of Euroimmun and Mt. Sinai would be above 35%, rendering over a third of positive serology test results as likely false positives.

The test by Roche pharmaceuticals, advertised as highly accurate,^36^, indeed has high PPV and low false positive rates even at 1%, with an upper limits for the false positive rate of 25.2%.The Roche test is the only test available by the end of May 2020 that could have reasonably claimed to expect more true positive results than false positive results in populations with low prevalence (1% or less). However, it would have still been reasonable to expect that up to one quarter of positive test results could have come from patients lacking antibodies to SARS-COV-2. Given the sample size^33^ of 29 for calculating sensitivity, the associated uncertainty could be compounded in the estimates of PPV and FPR, rendering the upper bounds critical to measure and understand. Even six months later, only one test, (Simens Atellica IgG) has improved on this figure, with the smallest upper confidence limit for FPR of 24.5% at 1% prevalence and a corresponding FPR point estimate of about 9%.

## 4 Application to Specific Locations

In this section, we evaluate PPV and FPR for a set of areas with three local studies of seroprevalence, in California, New York, and Boston. When rigorous prevalence estimates are unavailable, we use the proportion of positive tests as surrogates for prevalence for the purpose of estimating the rates of false positives in these studies. Rationale for and limitations of this approach along with a sensitivity analysis are discussed in the Supplementary Material.

### 4.1 Santa Clara County

An early seroprevalence study was conducted in Santa Clara County.However, after extensive scrutiny, the authors provided updated estimates. Based on the updated preprint^37^, the estimated prevalence adjusted for test performance characteristics were 1.2% (95%CI 0.7-1.8%) unweighted and 2.8% (95CI 1.3-4.7%) for weighted estimates based on demographic characteristics in Santa Clara County. Reanalyses^38^,^39^ reported updated seroprevalence ranges of 0.27% and 3.21%^38^ and 0% to 2.1%^39^. An estimate of prevalence in California from March 31 to April 7 is 0.9%^31^. Estimates of sensitivity and specificity vary^37–39^; we use the Bayesian posterior estimates^39^ combining information from all sources as 99.5% specificity with a 95% posterior interval of (98.8%,99.8%) and 81.8% sensitivity (64.2%.91.0%)

Predictive values in the Santa Clara Study nearly spanned the entire range of probabilities. Using prevalence values reflecting prior estimates ranging from 0% to 5%, the PPV in Santa Clara County at the time of the study ranges from 0% to 96% (Figure 5). Prevalence near the low but nonzero end of the updated estimates (e.g. 0.2%)^38^,^39^ correspond to PPV ranging from about 9.7% to to 47.7%, indicating that between about 26 and 46 of the 50 positive tests could be false. For prevalence near the high end of the updated estimates (4.7%)^37^ ranges from 73.0% to 95.8%, or 2 to 14 false positives. Reanalyses estimate smaller upper bounds on prevalence (2.1%^39^ and 3.2%^38^), which PPV ranges of 52.2% to 92.3% or 63.9% to 93.8%. These estimates correspond to false positive counts ranging from 2 to 24 or 3 to 18.

**Figure 5.**
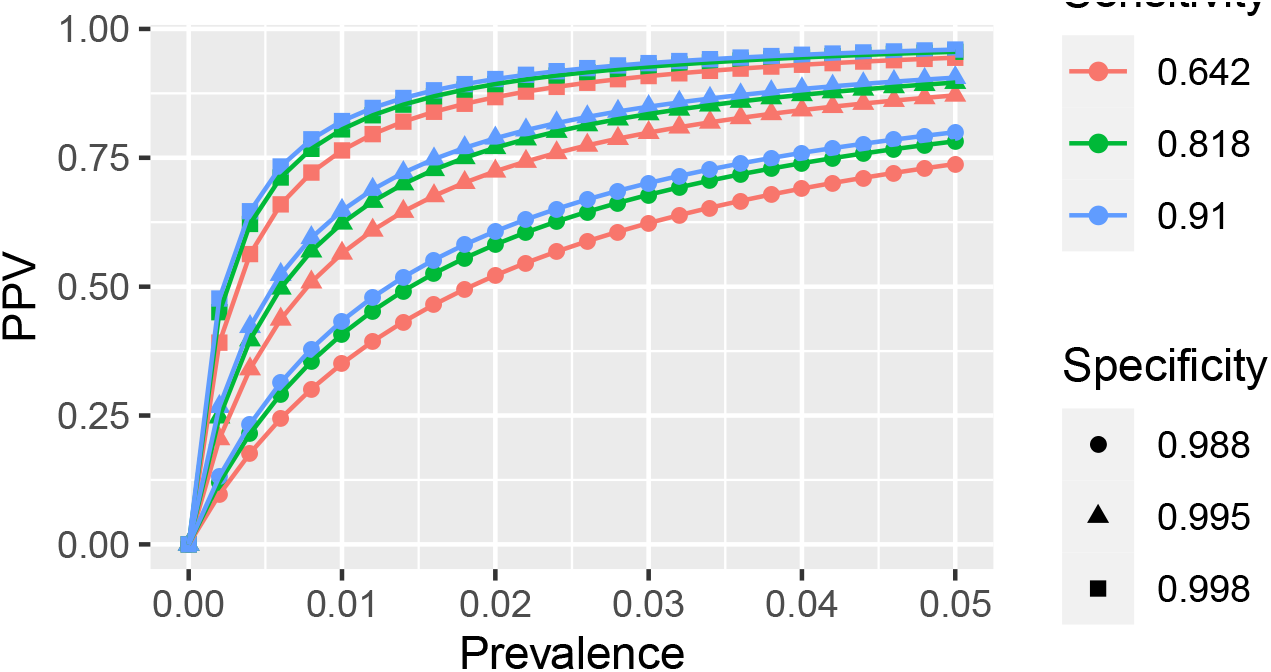
PPV for Antibodies test in Santa Clara County Study^37^. To reflect variation within and between references^37^–39, prevalence values are included from 0% to 5% by 0.2%.

### 4.2 New York

On April 23, Govornor Andrew Cuomo announced results from a serology study in New York^40^. Seropositivity rates were 13.9% for the state overall and differed by region. New York City, Long Island, Westchester and Rockland, and elsewhere in New York, respectively, had 21.2%, 16.7%, 11.7%, and 3.6% seropositive. Specificity for the test was reported to be in the range of 93-100%^41^, and sensitivity was not listed. However, the test was attributed to Wadsworth Center by the New York State Department of Health; the Wadsworth test parameters are reported^33^ in Table 1. Results were then updated^42^ on May 2. At that time, 12.3% of the population of New York state was reported to have Covid-19 antibodies based on a test of 15,000 people. By region, these figures ranged from 1.2% seropositive in North Country to 19.9% in New York City. We estimate the PPV for all combinations of values reported in all of these sources assuming the same serology tests were used in both studies. Figure 6 shows the range of PPV based on each of these possible values of sensitivity, specificity, and prevalence. New York City and Long Island had the highest prevalence and highest PPV, ranging from 74% to 98% and 60% to 97% in all scenarios. Statewide and other areas are in the middle: 60% to 96% for Rockland, 65% to 97% for statewide. By contrast, PPV can be as low as 30% for the rest of the state, assuming a prevalence 3.6%, if the specificity is 93% or as high as 87% if sensitivity is at the upper limit of the confidence interval reported in the EUA^33^. Even worse, areas with low prevalence^42^such as North Country (1.2%), Central NY (1.9%), and Capital District (2.2%) had PPV estimates ranging from 12% to 69%, 18% to 78% and 21% to 81% In other words, the false positive rate in New York ranged from 2% to 88% depending on the region and assumed prevalence under consideration and uncertainty in the sensitivity and specificity.

**Figure 6.**
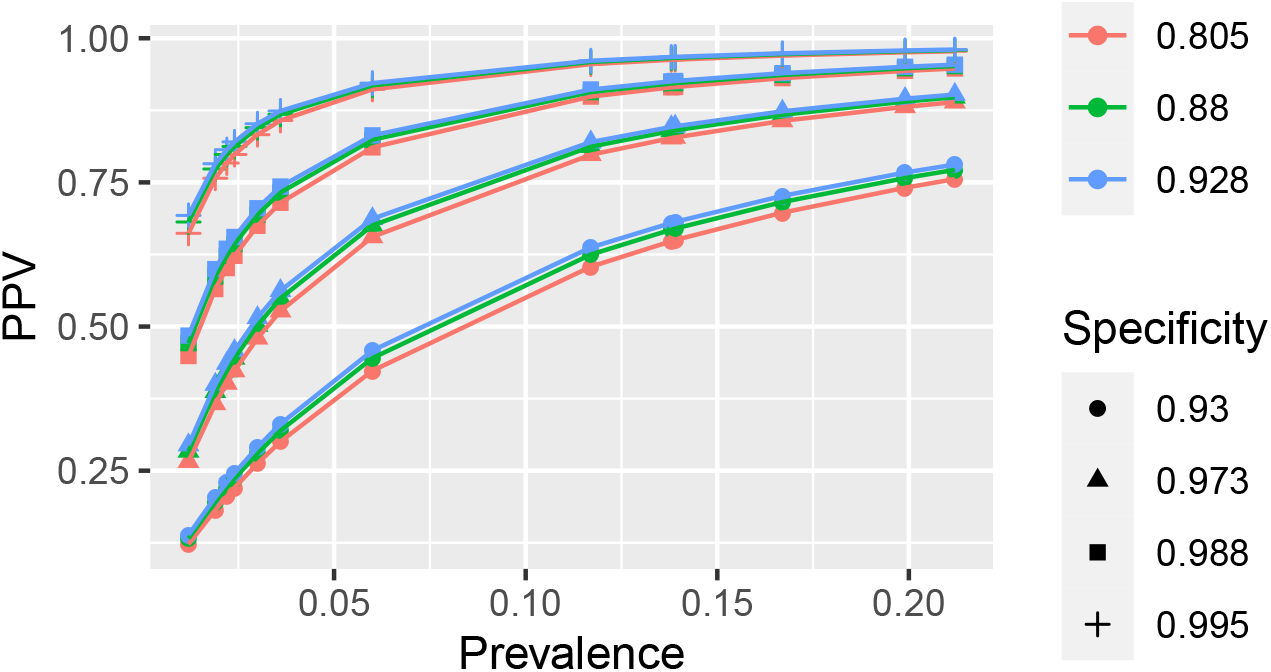
PPV for Antibodies test in New York Overall and by Region^33,40–42^

#### 4.2.1 Chelsea, Massachusetts

Researchers at Massachusetts General Hospital conducted a seroprevalence study in the city of Chelsea^22^ and reported that 31.5% of participants tested positive. The researchers on the Chelsea study reported specificity exceeding 99.5%. However, the manufacturer, BioMedomics, which is not part of the labs with EUA in Section 3.3, reports a sensitivity of 88.66% and specificity of 90.63%,^43^.

Assuming prevalence of 31.5% and specificity and sensitivity values reported by Biomedomics^43^, PPV was only about 81%. This means that in this sample of 63 positives, about 51 are expected to be true positives and about 12 are expected to be false positives. Thus, the prevalence estimate in Chelsea reported in the press based on this sample alone is likely to be too high. In addition, participants were recruited by a convenience sample of people outside on a particular street, which may not be representative of the general population in Chelsea.

## 5 Discussion

Antibodies tests can yield two possible errors with different implications^25^. Consequences of false negative test results would likely relate to failing to remove negative effects of limitations during the pandemic. For example, assuming that antibodies indeed confirm protection, then people with antibodies who test negative would be safe to return to work but their negative test might convince them to remain at home. This would prolong the negative mental and physical effects of social isolation as well as economic effects to individuals and society overall. Fortunately, the false negative rate was under 10% in all scenarios.

Unfortunately, the false positive rate can be shockingly high. Based on the prevalence estimated throughout the US and serology studies in California, New York and Boston, the FPR of antibody test results range from 2% to 88%. Point estimates of tests ests with an EUA^44^ reached 86% and upper limits reached 93% when the prevalence is 1%. Tests with low PPV and high FPR can be dangerous by giving patients with positive tests a false sense of security. Ironically, these people may then *increase* their risk of contracting Covid19 if they relax their use of protective measures, such as mask wearing and social distancing.

The timing of the test may impact the result, as discussed in the supplementary material. Briefly, seroconversion is the process during which antibodies develop after infected by Covid19 become detectable in the blood; the seroconversion duration could complicate the consideration of interpretation of serology test results. After infection, patients took about 3-40 days to develop detectable antibodies^45^,^46^. Typically after 14 days, most patients will develop antibodies. If the testing period is within 14 days, the sensitivities of the tests will be lower.

The number and implication of false positives is growing with large organizations encouraging widespread serology testing. Quest Diagnostics is offering tests by Abbott, Ortho-Clinical and Euroimmun for purchase^47^. The tests can have up to 86% FPR in locations with 1% prevalence. Even at 10% prevalence, over one-third of positives could be false. OneBlood, a non-profit blood donation and distribution organization, is encouraging large scale blood donation and then testing samples for antibodies using the OrthoClinical total test^48^. As discussed in section 3.3, the OrthoClinical test could have a false positive rate of over 50% with 1% prevalence or nearly 10% if the prevalence is 10%. Further, OneBlood is planning to use blood samples testing positive for antibodies as convalescent plasma. In fact, on August 23, the FDA granted EUA for convalescent plasma in patients hospitalized with Covid-19^49^. As shown in this paper, single serology tests of the general population in low prevalence areas could yield a large number of false positives, which could inadvertently harm patients. Using all samples that test positive could mean that large numbers of samples falsely considered to contain convalescent plasma which would become inadvertent placebos for patients actively struggling with Covid-19.

More recently, with the impending approval of vaccine candidates, there is a need to determine the prioritization of groups throughout the dose distribution process. In some discussions^50^, patients who previously contracted covid-19 could be considered lower priority for receiving vaccine until after the uninfected public. Given the false positives discussed in serology tests in this paper, it would be imprudent to determine past disease status by widespread serology testing alone. In this case, uninfected individuals who falsely test positive on serology tests would be denied the vaccine in a timely manner and therefore would have to wait longer for the opportunity to vaccinate and protect themselves from the virus. We recommend against using single serology tests to screen for prior infection. In fact, the length of protection from reinfection is unknown for individuals with prior exposure either to the disease itself or via vaccination.

One recommendation for individuals who test positive is to consider a follow-up antibody test^51^. For instance, if the FPR is 50%, assuming independent tests, the probability of two false positives drops in half (25%). The FDA includes a calculator for PPV of individual and combined tests^33^. Follow-up tests are common in other diseases with low PPV screening methods, such as mammograms for breast cancer^52^. For instance, one could use a highly sensitive test with sensitivity/specificity, say, 99% and 90 %, respectively, as the initial test, and then followed up a highly specific test with sensitivity/specificity of 90% and 99% respectively, as the second test. Then, the FPR (i.e.,1-PPV) would drop from 81.9% to 10% for a place with prevalence at 1%. For a place with high prevalence at 10%, the FPR could drop from 47.6% to 1% Another idea is to test all contacts for antibodies and use their results as evidence to support or refute the original serology test. Pursuing contacts of additional seropositive individuals may increased contact tracing and testing, which can either hinder growth of future outbreaks or divert scarce resources from higher risk contacts^25^. Moreover, increased testing brings cost and feasibility concerns.

There are some additional limitations of our paper. For instance, some of the information may become outdated quickly. Tests operating under an FDA EUA will likely increase over time. For instance, Abbott Alinity was added to the original 12 tests made available by FDA under EUA in early May 2020 while writing the first draft of this paper. At the time, the FDA noted that at least 160 serology tests were available before the FDA increased its oversight^53^. This implies some of the antibody tests on the market might even have lower sensitivity or specificity than those included in this paper and therefore have even higher FPR. Indeed, between the original paper submission in June 2020 and this revision in December 2020, two tests, AutoBio and ChemBio, had their FDA EUA revoked^54^,^55^. Notably, valaues for BioRad and Ortho-Clinical changed, and Diasorin added an IgM test along with its earlier IgG test. Similarly, if an infected patient takes the test before antibodies are developed, then the sensitivity will be lower. An extended discussion is included in the Supplementary Material. At the same time, prevalence may increase over time at least for some of the cities or towns. A reference to prevalence estimates calculated by the CDC and current as of the writing of this revision is provided^32^. Importantly, neither the results nor the interpretation for serology tests generalize to diagnostic tests. The Supplementary Material provides a brief discussion.

In conclusion, serology tests for the novel coronavirus generally have low false negative rates and highly volatile false positive rates. While false positive rates decrease with increasing prevalence, current prevalence estimates remain low in most areas of the US. With increasing serology testing and likely increased reporting of testing results, it is critical to understand these values and interpret test results properly. We hope that this context and interpretation can aid doctors, patients, researchers, and policy makers in informed decision making, which may even save lives.

## 6 Methods

We collected reported sensitivity and specificity values of serology tests with EUA approval by the US FDA. Prevalence estimates were also collected to determine an appropriate range for the plots. These values were combined to produce estimates of PPV and NPV for a variety of input parameter values. We then honed in on specific FDA tests and areas that have conducted serology tests to provide estimates and uncertainty for PPV and false positive rates.

All programs utilized R version 3.6.1^56^. PPV and NPV were calculated using package MKmisc^57^. Plots were created with packages plotly^58^ and ggplot2^59^. Figures 1 and 2, as higher dimensional plots, were designed to allow interactive visualization. Code to generate the plots and view them in an interactive mode may be downloaded from our github repository at https://github.com/nbrownst/AntibodiesPredictiveValues.

## Data Availability

Data and code utilized for this paper are available in our github repository at https://github.com/nbrownst/AntibodiesPredictiveValues.

## 7 Acknowledgements

The authors ackowledge support from the National Cancer Institute (5P30 CA076292-22) and the Biostatistics and Bioinformatics Core at the H. Lee Moffitt Cancer Center & Research Institute.

## References

1. Pooladanda, V., Thatikonda, S. & Godugu, C. The current understanding and potential therapeutic options to combat covid-19. Life Sci. 117765 (2020).

2. Borges do Nascimento, I.J. et al. Novel coronavirus infection (covid-19) in humans: a scoping review and meta-analysis. J. clinical medicine 9, 941 (2020).

3. do Nascimento, I. J. B. et al. Coronavirus disease (covid-19) pandemic: An overview of systematic reviews. medRxiv (2020).

4. 9. Rajkumar, R. P. Covid-19 and mental health: A review of the existing literature. Asian J. Psychiatry 102066 (2020).

5. Barua, S. et al. Understanding coronanomics: The economic implications of the coronavirus (covid-19) pandemic. Manuscript (2020).

6. Fernandes, N. Economic effects of coronavirus outbreak (covid-19) on the world economy. Available at SSRN 3557504 (2020).

7. Ozili, P. K. & Arun, T. Spillover of covid-19: impact on the global economy. Available at SSRN 3562570 (2020).

8. Di Gennaro, F. et al. Coronavirus diseases (covid-19) current status and future perspectives: A narrative review. Int. J. Environ. Res. Public Heal. 17, 2690 (2020).

9. Patrick, S. L. & Cormier, H. C. Are our lives the experiment? covid-19 lessons during a chaotic natural experiment a commentary. Heal. Behav. Policy Rev. 7, 165–169, DOI: doi:10.14485/HBPR.7.2.10 (2020).

10. Chen, W.-H., Strych, U., Hotez, P. J. & Bottazzi, M. E. The sars-cov-2 vaccine pipeline: an overview. Curr. tropical medicine reports 1–4 (2020).

11. Prachar, M. et al. Covid-19 vaccine candidates: Prediction and validation of 174 sars-cov-2 epitopes. bioRxiv (2020).

12. Eyal, N., Lipsitch, M. & Smith, P. G. Human challenge studies to accelerate coronavirus vaccine licensure. The J. Infect. Dis. (2020).

13. Spinney, L. When will a coronavirus vaccine be ready. The Guard. Retrieved 18 (2020).

14. Omer, S. B., Malani, P. & Del Rio, C. The covid-19 pandemic in the us: a clinical update. JAMA (2020).

15. Velavan, T. P. & Meyer, C. G. The covid-19 epidemic. Trop. medicine & international health 25, 278 (2020).

16. Emanuel, E. J. et al. Fair allocation of scarce medical resources in the time of covid-19. New Engl. J. Medicine 0, ull, DOI: 10.1056/NEJMsb2005114 (0). https://doi.org/10.1056/NEJMsb2005114.

17. John, B. S. Life in the time of covid-19: a crisis and a crocus. https://www.uchealth.org/today/life-in-the-time-of-covid-19-a-crisis-and-a-crocus/. Accessed: 2020-05-05.

18. Winter, A. K. & Hegde, S. T. The important role of serology for covid-19 control. The Lancet. Infect. Dis. (2020).

19. World Health Organization and others. “immunity passports” in the context of covid-19: scientific brief, 24 april 2020. Tech. Rep., World Health Organization (2020).

20. Farnsworth, C. W. & Anderson, N. W. Sars-cov-2 serology: Much hype, little data. Clin. Chem. (2020).

21. Torres, R. & Rinder, H. M. Double-edged spike: Are sars-cov-2 serologic tests safe right now? Am. J. Clin. Pathol. (2020).

22. Vogel, G. First antibody surveys draw fire for quality, bias. Science 368, 350–351, DOI: 10.1126/science.368.6489.350 (2020). https://science.sciencemag.org/content/368/6489/350.full.pdf.

23. Phelan, A. L. Covid-19 immunity passports and vaccination certificates: scientific, equitable, and legal challenges. Lancet (London, England) (2020).

24. World Health Organization. Population-based age-stratified seroepidemiological investigation protocol for covid-19 virus infection. Tech. Rep., World Health Organization (2020). Available at: https://www.who.int/publications-detail/population-based-age-stratified-seroepidemiological-investigation-protocol-for-covid-19-virus-infection.

25. Gronvall, G. et al. Developing a national strategy for serology (antibody testing) in the united states. Tech. Rep., Johns Hopkins Center for Health Security (2020). Available at: https://www.centerforhealthsecurity.org/our-work/publications/developing-a-national-strategy-for-serology-antibody-testing-in-the-US.

26. Pepe, M. S. et al. The statistical evaluation of medical tests for classification and prediction (Medicine, 2003).

27. Fleiss, J. L., Levin, B. & Paik, M. C. Statistical methods for rates and proportions (john wiley & sons, 2013).

28. Bonislawski, A. False positives could undermine utility of sars-cov-2 serology testing. https://www.360dx.com/infectious-disease/false-positives-could-undermine-utility-sars-cov-2-serology-testing (2020). [Online; accessed 13-May-2020].

29. Lu, F. S., Nguyen, A. T., Link, N. & Santillana, M. Estimating the prevalence of covid-19 in the united states: Three complementary approaches. medRxiv DOI: 10.1101/2020.04.18.20070821 (2020). https://www.medrxiv.org/content/early/2020/04/23/2020.04.18.20070821.full.pdf.

30. Breton, T. R. An estimate of unidentified and total us coronavirus cases by state on april 11, 2020. Available at SSRN 3583941 (2020).

31. Benatia, D., Godefroy, R. & Lewis, J. Estimating covid-19 prevalence in the united states: A sample selection model approach. medRxiv (2020).

32. Cdc covid data tracker. https://covid.cdc.gov/covid-data-tracker/#national-lab (2020). Accessed: 2020-11-25.

33. Eua authorized serology test performance. https://www.fda.gov/medical-devices/emergency-situations-medical-devices/eua-authorized-serology-test-performance (2020). Online: Accessed 05-23-2020.

34. Eua authorized serology test performance. https://www.fda.gov/medical-devices/emergency-situations-medical-devices/eua-authorized-serology-test-performance (2020). Online: Accessed 11-23-2020.

35. Arya, R., Antonisamy, B. & Kumar, S. Sample size estimation in prevalence studies. The Indian J. Pediatr. 79, 1482–1488 (2012).

36. Elecsysä anti-sars-cov-2. https://diagnostics.roche.com/us/en/products/params/elecsys-anti-sars-cov-2.html#productInfo. [Online; accessed 5-May-2020].

37. Bendavid, E. et al. Covid-19 antibody seroprevalence in santa clara county, california. medRxiv DOI: 10.1101/2020.04.14.20062463 (2020). https://www.medrxiv.org/content/10.1101/2020.04.14.20062463v2.full.pdf.

38. Bennett, S. T. & Steyvers, M. Estimating covid-19 antibody seroprevalence in santa clara county, california. a re-analysis of bendavid et al. medRxiv (2020).

39. Gelman, A. & Carpenter, B. Bayesian analysis of tests with unknown specificity and sensitivity. http://www.stat.columbia.edu/gelman/research/unpublished/specificity.pdf (2020). Accessed 05-25-2020.

40. Video, audio, photos & rush transcript: Amid ongoing covid-19 pandemic, governor cuomo announces state health department will partner with attorney general james to investigate nursing home violations. https://www.governor.ny.gov/news/video-audio-photos-rush-transcript-amid-ongoing-covid-19-pandemic-governor-cuomo-announces-12 (2020). Online: Accessed 05-15-2020.

41. Bonislawski, A. New york, california serology studies give early estimates of covid-19 prevalence. https://www.360dx.com/infectious-disease/new-york-california-serology-studies-give-early-estimates-covid-19-prevalence (2020). [Online; Accessed 13-May-2020].

42. Amid ongoing covid-19 pandemic, governor cuomo announces results of completed antibody testing study of 15,000 people showing 12.3 percent of population has covid-19 antibodies. https://www.governor.ny.gov/news/amid-ongoing-covid-19-pandemic-governor-cuomo-announces-results-completed-antibody-testing (2020). Online: Accessed 05-16-2020.

43. Covid-19 igm/igg rapid test. https://biomedomics.com/products/infectious-disease/covid-19-rt/. [Online; accessed 13-May-2020].

44. U.S. Food and Drug Administration. Policy for coronavirus disease-2019 tests during the public health emergeyncy (revised). Tech. Rep., U.S. Department of Health and Human Services, Food and Drug Administration (2020). Available at: https://www.fda.gov/media/135659/download.

45. Xiang, F. et al. Antibody detection and dynamic characteristics in patients with covid-19. Clin. Infect. Dis. (2020).

46. Flodgren, G. Immunity after sars-cov-2 infection, 1st update–. Lancet Infect. Dis. 23, 23 (2020).

47. Patient and healthcare provider fact sheets for covid–19 testing. https://www.questdiagnostics.com/home/Covid-19/factsheet/ (2020). Accessed 05-25-2020.

48. Covid-19 antibody testing. https://www.oneblood.org/lp/oneblood-covid-19.stml. Online: Accessed 05-23-2020.

49. Fda issues emergency use authorization for convalescent plasma as potential promising covid–19 treatment, another achievement in administration’s fight against pandemic. https://www.fda.gov/news-events/press-announcements/fda-issues-emergency-use-authorization-convalescent-plasma-potential-promising-covid-19-treatment. Online; accessed 7-Dec-2020.

50. Wang, W. et al. Global, regional, and national estimates of target population sizes for covid-19 vaccination. medRxiv (2020).

51. Yi, G., He, W., Lin, D.K.-J. & Yu, C.-M. Covid-19: Should we test everyone? (2020). 2004.01252.

52. Nelson, H. D. et al. Screening for breast cancer: an update for the us preventive services task force. Annals internal medicine 151, 727–737 (2009).

53. fda steps up scrutiny coronavirus antibody tests ensure accuracy. https://www.washingtonpost.com/health/2020/05/04/fda-steps-up-scrutiny-coronavirus-antibody-tests-ensure-accuracy/. Online; accessed 4-May-2020.

54. Hinton, D.M. Re: Revocation of eua200349. https://www.fda.gov/media/140908/download (2020). Accessed: 2020-12-7.

55. “coronavirus (covid-19) update: Fda revokes emergency use authorization for chembio antibody test”. https://www.fda.gov/news-events/press-announcements/coronavirus-covid-19-update-fda-revokes-emergency-use-authorization-chembio-antibody-test (2020). Accessed: 2020-12-07.

56. R Core Team. R: A Language and Environment for Statistical Computing. R Foundation for Statistical Computing, Vienna, Austria (2019).

57. Kohl, M. MKmisc: Miscellaneous functions from M. Kohl (2019). R package version 1.6.

58. Sievert, C. Interactive Web-Based Data Visualization with R, plotly, and shiny (Chapman and Hall/CRC, 2020).

59. Wickham, H. ggplot2: Elegant Graphics for Data Analysis (Springer-Verlag New York, 2016).

