## Appendices for "Predictive values, uncertainty, and interpretation of serology tests for the novel coronavirus"

### Supplementary Material for “Are antibodies tests accurate? Understanding predictive values and uncertainty of serology tests for the novel coronavirus”

**Naomi C Brownstein[1,*] and Yian Ann Chen[1]**

[1]Moffitt Cancer Center, Department of Biostatistics and Bioinformatics, Tampa, FL, USA
[*]

#### ABSTRACT

Supplementary material is included in this document.

### 1 Introduction

We include four appendices. Section 2 reviews Effect of Time from Infection on antibodies test results and includes a analysis of one of the FDA tests for individuals tested too early, i.e. less than two weeks after initial infection. Section 3 and 4 includes intermediate calculations and commentary on the difference between the test positivity rate and estimated prevalence. Section 4.3 briefly mentions differences in interpretation in these statistics between serology and diagnostic tests. Section 5 includes additional tests approved for EUA between the paper submission in spring of 2020 and the revision submission in Fall of 2020.

### 2 Effect of Time from Infection

We provide an example of the effect of test timing on PPV. As noted in section 3.3 of the main text, the test by Roche pharmaceuticals[1], reported relatively high PPV compared to its competitors (Table 1and Figure 4, with 95% confidence intervals of (88%,100%) for specificity and (99.61%, 99.91%) for sensitivity when the test is taken at least 14 days after infection. However, when the test is given early, i.e., within 6 days, the sensitivity is only about 65.5 % (56.1 – 74.1 %). When the test is given between 7-13 days, the sensitivity is 88.1 % (77.1 – 95.1 %). Figure S1 displays the PPV by prevalence.

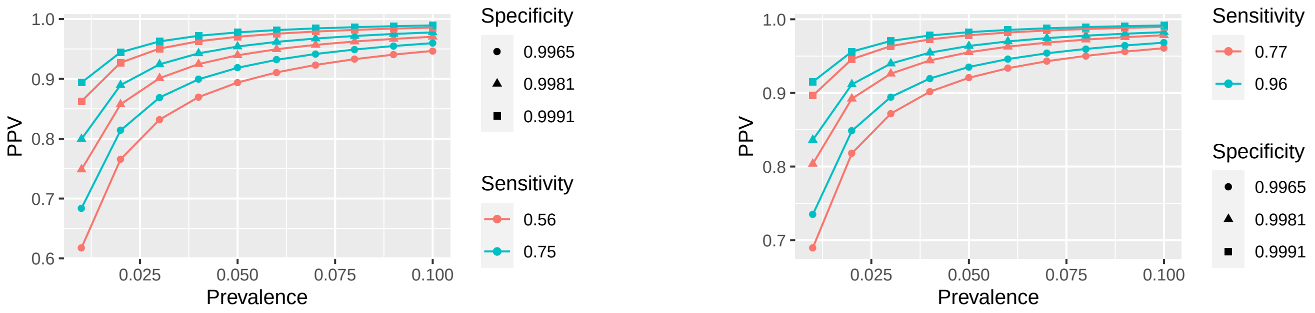


**(a)** 0-6 days

**(b)** 7-13 days

**Figure S1.** PPV by prevalence for Roche test taken prior to 2 weeks post infection

### 3 Definitions and equations

For the illustrative purposes of this paper, we include simplifying assumptions. First, we assume that people who have not yet had a SARS-Cov-2 infection (with or without symptoms) should lack antibodies. Conversely, we assume that a person who does not have antibodies either has not yet been infected with SARS-CoV-2 or has been infected too recently for the body to have developed antibodies. Limitations and further discussion on these items are included in 2 and the discussion section of the main text, as well as in external sources[2,3] .

Statistical notation for these definitions follow. For a randomly selected person, consider the following random variables. $D$ is the indicator of whether the person was previously infected with novel coronavirus disease (SARS-CoV-2) and has antibodies. $T$ is the indicator of if the antibodies test result is positive. The prevalence of SARS-Cov-2 infection and antibodies posession is $p = p(D = 1)$. PPV and NPV are defined by equations (3) and (4):

$$sensitivity = P(T = 1|D = 1) \tag{1}$$

$$specificity = P(T = 0|D = 0) \tag{2}$$

$$PPV = P(D = 1|T = 1) \tag{3}$$

$$NPV = P(D = 0|T = 0) \tag{4}$$

One can invoke Bayes Rule[4] and the law of total probability[5] to show that:

$$P(T=1) = p*sensitivity + (1-p)*(1-specificity) \tag{5}$$

$$PPV = \frac{p*sensitivity}{p*sensitivity + (1-p)*(1-specificity)} \tag{6}$$

$$NPV = \frac{(1-p)*specificity}{(1-p)*specificity + p*(1-sensitivity)} \tag{7}$$

Although sensitivity and specificity clearly differ from PPV and NPV, respectively, these distinct quantities are often misinterpreted in practice[6,7]. Importantly, the PPV and NPV of serology tests depend on pretest parameters and on the prevalence of SARS-CoV-2, which is unknown and difficult to measure[8]. PPV and NPV correspond to probabilities that test results of each type are correctly classified. Their complements refer to false testing ratse. False positives refer to positive serology tests for patients lacking antibodies, while false negatives refer to negative tests for patients with antibodies.

$$FPR = 1 - PPV = P(D=0|T=1) \tag{8}$$

$$FNR = 1 - NPV = P(D=1|T=0) \tag{9}$$

#### 4 Additional Calculations and Application to Serology Studies

The studies in New York and Chelsea provided only positive testing rates, not prevalence estimates. The positive testing rate is not the same as the prevalence. We conduct sensitivity analyses after including initial calculations. Results were generally similar. Often the estimated prevalence (and PPV) was slightly lower than the seropositivity rate.

First, we break down the probability of testing positive in equation (10).

$$\begin{aligned} P(T=1) &= P(T=1 \cap D=1) + P(T=1 \cap D=0) \\ &= P(T=1|D=1)P(D=1) + P(T=1|D=0)P(D=0) \\ &= P(T=1|D=1)P(D=1) + [1-P(T=0|D=0)][1-P(D=1)] \\ &= p*sensitivity + (1-p)*(1-specificity) \\ &= p*(sensitivity + specificity - 1) + (1-specificity) \end{aligned} \tag{10}$$

The proportion testing positive in a seroprevalence study can be used to estimate the true prevalence. Denote $\hat{p}_t$ as the observed proportion who have have antibodies. We can substitute $\hat{p}_t$ on the left hand side of (10) as an estimate for $P(T=1)$ and solve for the prevalence $\hat{p}$. The result is equation (11):

$$\begin{aligned} \hat{p} &= \frac{\hat{p}_t - (1-specificity)}{sensitivity + specificity - 1} \\ &= \frac{specificity + \hat{p}_t - 1}{sensitivity + specificity - 1} \\ &= \frac{specificity - (1-\hat{p}_t)}{sensitivity + specificity - 1} \end{aligned} \tag{11}$$

It is important to note that this means not every combination of sensitivity and specificity is possible for a given positive testing rate, as otherwise the estimated prevalence will be negative[9]. Given that sensitivity and specificity are usually large (e.g. each exceeding 80%), we would expect

$$sensitivity + specificity - 1 > 0$$

In order for the prevalence estimate to be non-negative, this means that the numerator must also be non-negative, and we would expect

$$\hat{p}_t \geq (1-specificity) \tag{12}$$

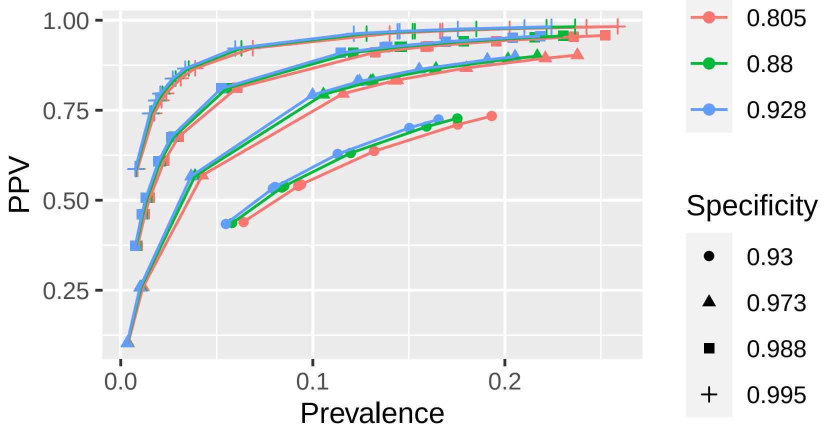


**Figure S2.** Sensitivity Analysis of PPV by prevalence in New York calculated by Equation (11)

That is, the positive rates observed in a seroprevalence study, $\hat{p}_t$ should be at least be large as the false positive rate of the antibody test used in that study. Indeed, if we saw fewer positive tests than the proportion of (false) positive tests expected by chance if everyone were lacking antibodies, then we would have little evidence to suggest that the prevalence is nominally different from zero. A sensitivity analysis follows for the studies in Chelsea and New York, which reported only seropositivity estimates. While the seropositive rates are not necessarily identical to the prevalence estimate, the estimated seropositivity rate was generally either close or a slight overestimate of the prevalence.

##### 4.1 Chelsea

In Chelsea, the original seropositivity rate was 31.5%. The updated prevalence estimates based on (11) was 27.9%, which is about 3.6% lower than the proportion who tested positive. Using this lower prevalence yields a lower PPV or 78.6%, which would correspond to about to expecting about 49 of the positive tests to be true positives and 14 to be false positives.

##### 4.2 New York

In the New York serology study presented in Section 4 of the main text, there were 156 estimates of PPV, resulting from the product of 3 possible values for sensitivity, 4 potential values for specificity, and 13 potential seropositivity estimates from the two studies[10,11].

Prevalence estimates were calculated based on (11). Unlike in the sensitivity analysis for Chelsea, 36 scenarios resulted in negative estimated prevalence values. For these combinations, the estimated PPV would be zero, which is lower than the PPV in the paper. The false positive rate in the sensitivity analysis would be even higher in the sensitivity analysis than in the paper.

For the 120 remaining scenarios with estimated positive prevalence values, the difference between the prevalence and positivity rate was generally small and centered close to zero. Among the 120 scenarios, the differences between the two estimated proportions ranged from -6.2% to 4.7% with a median difference of -0.3% and a mean difference of 0.6%. While the distribution of differences in prevalence was approximately symmetric, the distribution of differences in PPV was strongly left-skewed. The median difference in PPV was -0.96% and mean difference was -5.44%, with a range from -41% to 1.4%. The large differences in absolute value of PPV correspond to values where the PPV in this sensitivity analysis is much lower than the PPV when using seropositivity to estimate prevalence. However, most differences were small in absolute value, meaning that most PPV estimates were similar regardless of which value was used for prevalence.

An updated plot of PPV by prevalence estimated with (11) is shown in Figure S2. The shape is similar to Figure 6 in the main text, with noticeable missing segments for combinations with the lower bound of specificity and prevalence estimates outside of the range from about 5% to 20%. Small prevalence and specificity estimates likely violate Expression (12).

##### 4.3 Comparison with Other Types of Tests for SARS-Cov-2

It is important to note the difference in analysis and interpretation in this paper, compared to other tests for SARS-COV-2. Our paper showed that NPV was reasonably high and PPV was low for serology tests, with a lot of potential for harm for false positive serology tests, such as increasing risk for Covid19.

The interpretations differ for diagnostic tests. False positives for diagnostic tests would mean that an uninfected patient would be quarantined and their contacts tested. False negative diagnostic tests would mean that an infected person could be erroneously cleared, their future contacts consequently put at risk of exposure, and their past contacts less likely to be tested.

Thus, the potential harm for a false negative diagnostic test likely exceeds the potential harm for a false positive diagnostic test. Consequently, calculations and interpretations related to NPV should be emphasized for diagnostic tests to mitigate this harm.

While a rigorous analysis for other types are outside of the scope of this paper, we will provide some comments. We showed that PPV was closely related to specificity. Similarly, NPV is related closely to sensitivity. Thus, it is imperative that diagnostic tests for SARS-Cov-2 have high sensitivity. It is unclear whether this is true in practice, as there have been reports and analyses of diagnostic tests having high false negatives[12].

#### 5 Additional Results for Tests Approved for EUA after May 2020

The tests in this section were approved after the paper was originally submitted in June 2020 and added when responding to reviewer comments in early December 2020. For completeness, graphs of PPV by prevalence are included for all of these new tests to parallel the graphs of the tests available previously.

Results are found in Figures S3 to S9.

#### References


1. Elecsys® anti-sars-cov-2. https://diagnostics.roche.com/us/en/products/params/elecsys-anti-sars-cov-2.html#productInfo. [Online; accessed 5-May-2020].
2. Flodgren, G. Immunity after sars-cov-2 infection, 1st update–. Lancet Infect. Dis. **23**, 23 (2020).
3. Gronvall, G. et al. Developing a national strategy for serology (antibody testing) in the united states. Tech. Rep., Johns Hopkins Center for Health Security (2020). Available at: https://www.centerforhealthsecurity.org/our-work/publications/developing-a-national-strategy-for-serology-antibody-testing-in-the-US.
4. Westbury, C. Bayes' rule for clinicians: An introduction. Front. Psychol. **1**, 192, DOI: 10.3389/fpsyg.2010.00192 (2010).
5. Weisstein, E. W. Total probability theorem. https://mathworld.wolfram.com/TotalProbabilityTheorem.html. Online: Accessed 05-16-2020.
6. Gigerenzer, G., Gaissmaier, W., Kurz-Milcke, E., Schwartz, L. M. & Woloshin, S. Helping doctors and patients make sense of health statistics. Psychol. science public interest **8**, 53–96 (2007).
7. Trevethan, R. Sensitivity, specificity, and predictive values: Foundations, pliabilities, and pitfalls in research and practice. Front. Public Heal. **5**, 307, DOI: 10.3389/fpubh.2017.00307 (2017).
8. Lu, F. S., Nguyen, A. T., Link, N. & Santillana, M. Estimating the prevalence of covid-19 in the united states: Three complementary approaches. medRxiv DOI: 10.1101/2020.04.18.20070821 (2020). https://www.medrxiv.org/content/early/2020/04/23/2020.04.18.20070821.full.pdf.
9. Gelman, A. & Carpenter, B. Bayesian analysis of tests with unknown specificity and sensitivity. http://www.stat.columbia.edu/ gelman/research/unpublished/specificity.pdf (2020). Accessed 05-25-2020.
10. Video, audio, photos & rush transcript: Amid ongoing covid-19 pandemic, governor cuomo announces state health department will partner with attorney general james to investigate nursing home violations. https://www.governor.ny.gov/news/video-audio-photos-rush-transcript-amid-ongoing-covid-19-pandemic-governor-cuomo-announces-12 (2020). Online: Accessed 05-15-2020.
11. Amid ongoing covid-19 pandemic, governor cuomo announces results of completed antibody testing study of 15,000 people showing 12.3 percent of population has covid-19 antibodies. https://www.governor.ny.gov/news/amid-ongoing-covid-19-pandemic-governor-cuomo-announces-results-completed-antibody-testing (2020). Online: Accessed 05-16-2020.
12. Yi, G., He, W., Lin, D. K.-J. & Yu, C.-M. Covid-19: Should we test everyone? (2020). 2004.01252.

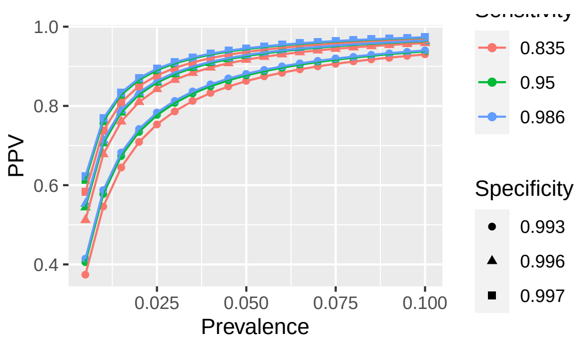


**(a)** Abbott-AdvDxAl

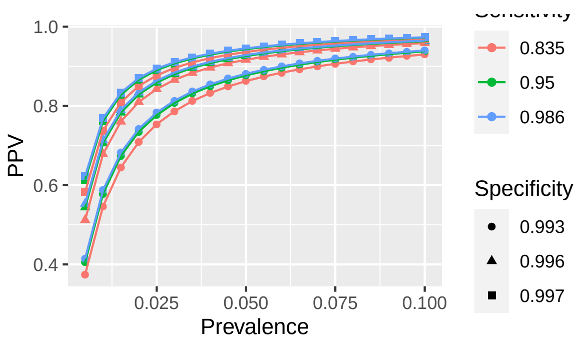


**(b)** Abbott-AdviseDxAr

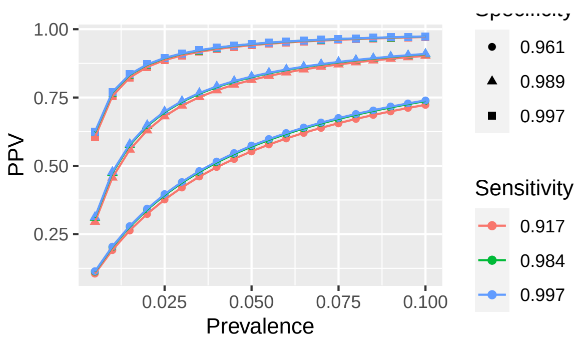


**(c)** AccessBio

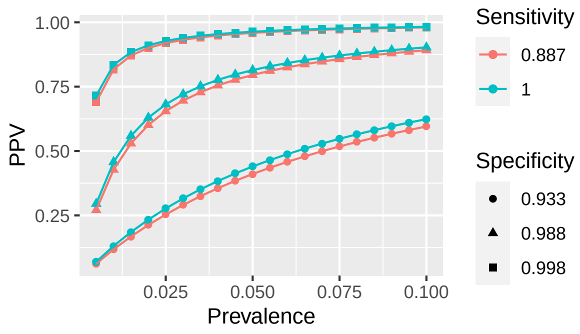


**(d)** AssureTech

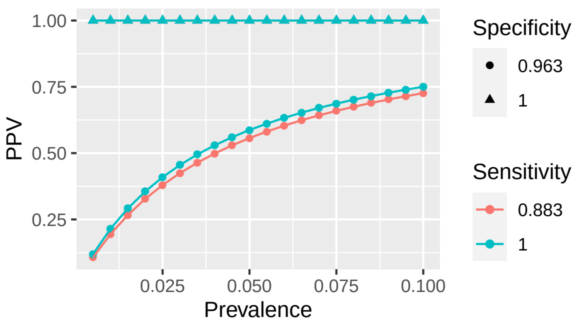


**(e)** Babson

**Figure S3.** EUA Tests: A-Ba

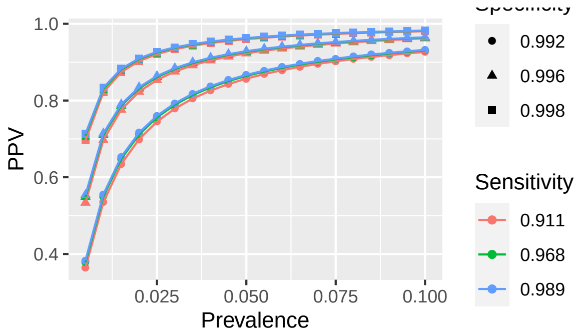


**(a)** BeckmanCoulter

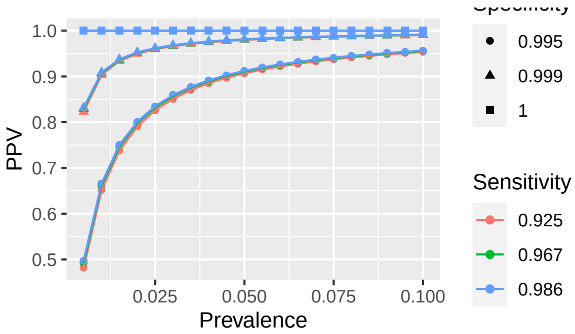


**(b)** BeckmanCoulter

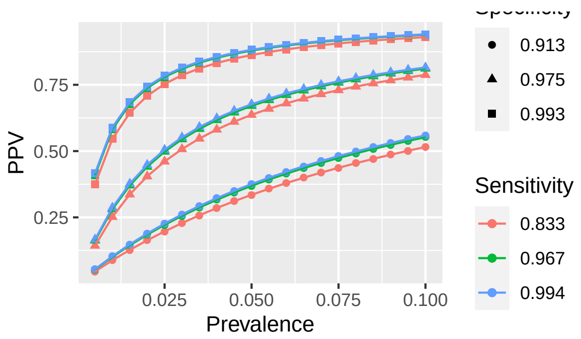


**(c)** Beijing Wantai ELISA

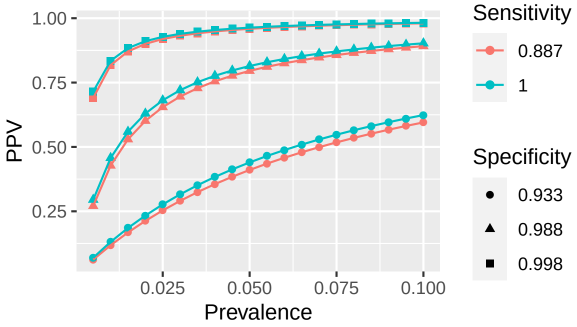


**(d)** Beijing Wantai Rapid

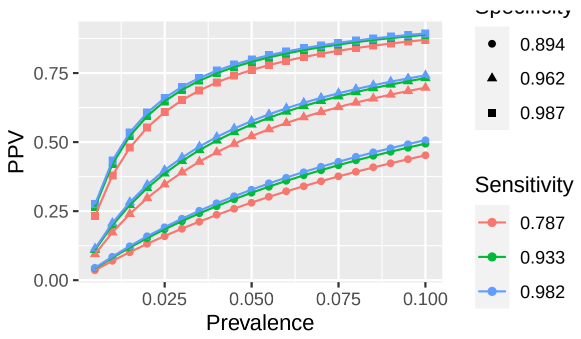


**(e)** BioCan

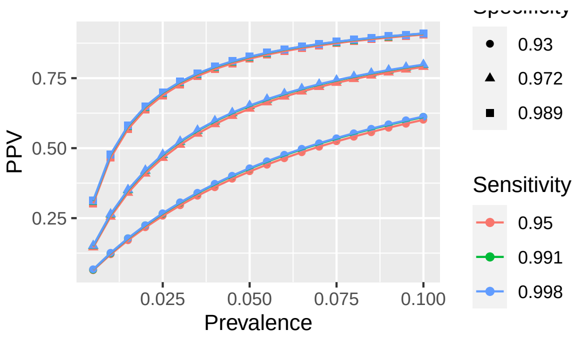


**(f)** BioCheck

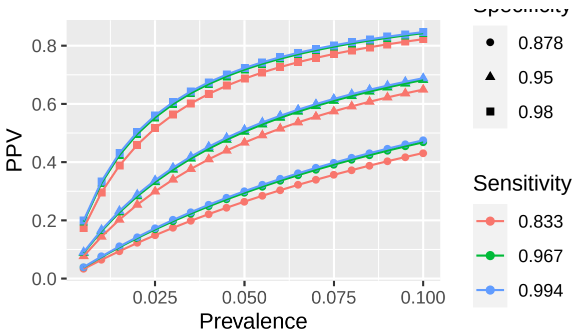


**(g)** Biohit

**Figure S4.** EUA Tests: Be-Bioh

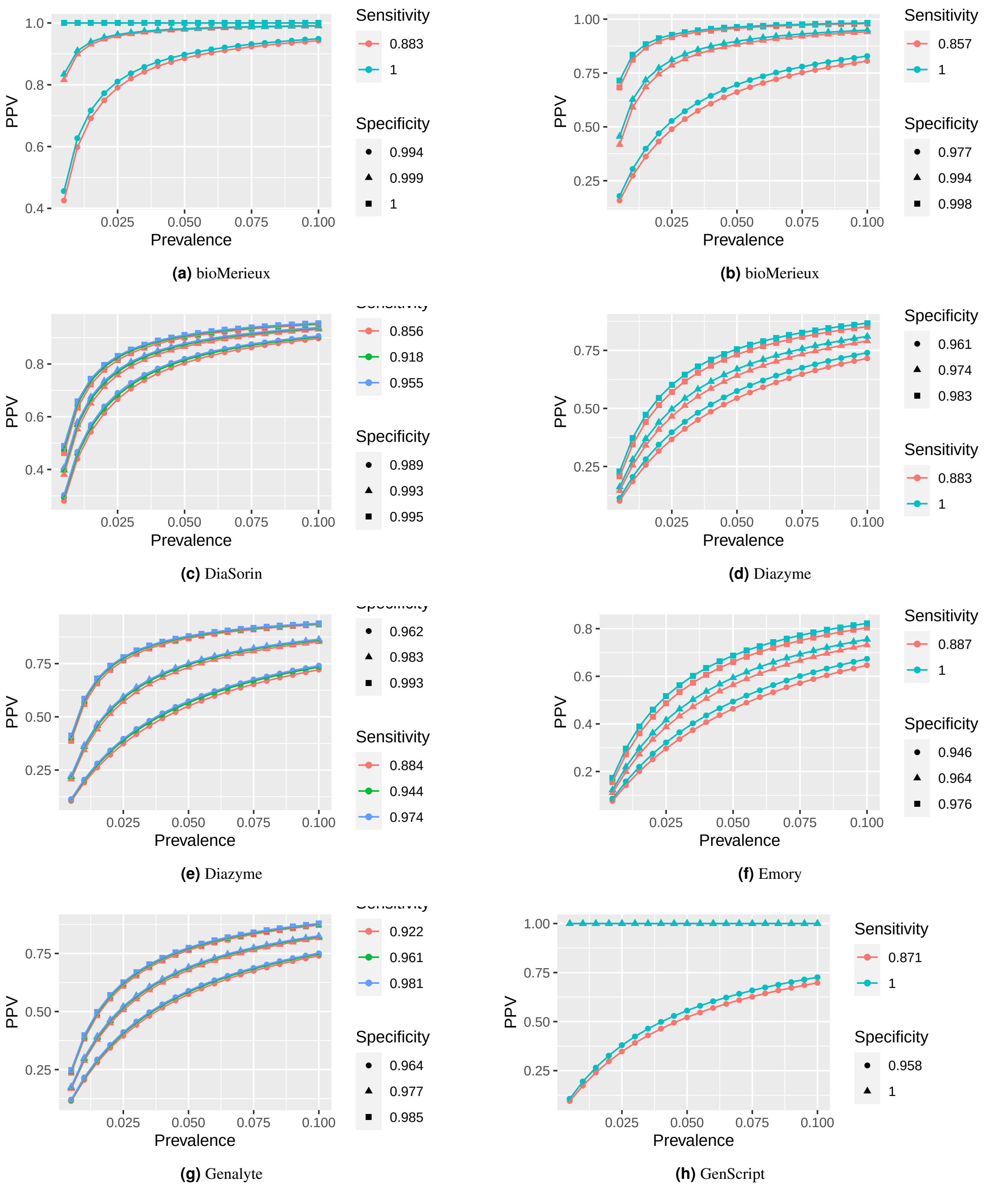


**(a)** bioMerieux **(b)** bioMerieux

**(c)** DiaSorin **(d)** Diazyme

**(e)** Diazyme **(f)** Emory

**(g)** Genalyte **(h)** GenScript

**Figure S5.** EUA Tests: BioM-G

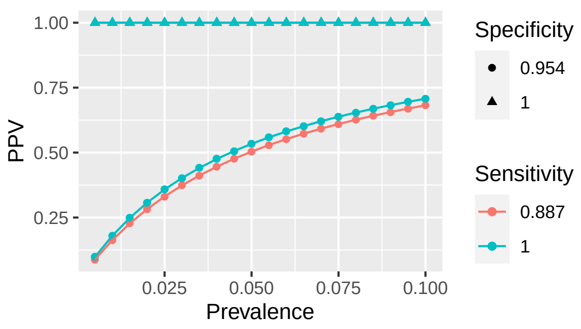


**(a)** HangzhouRapid

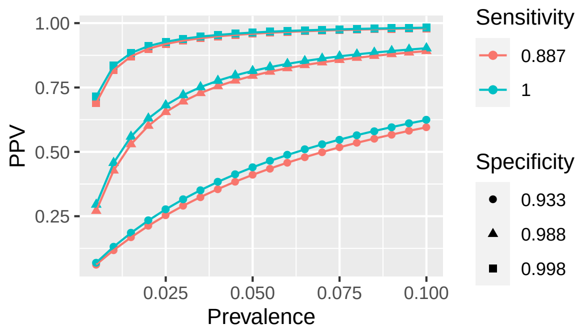


**(b)** HangzhouLaihe

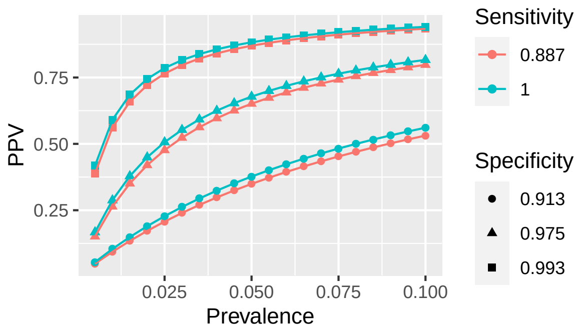


**(c)** Healgen

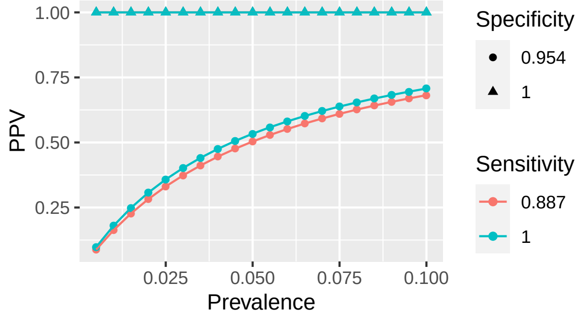


**(d)** InBios

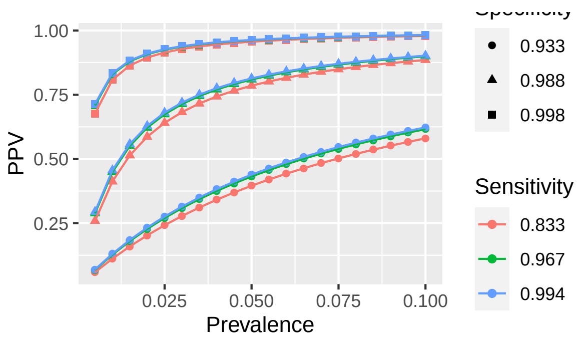


**(e)** InBios

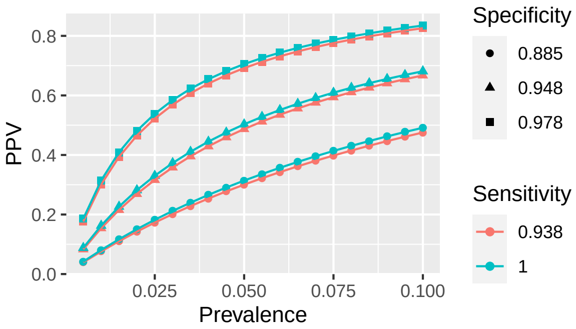


**(f)** JiangSu

**Figure S6.** EUA Tests: H-J

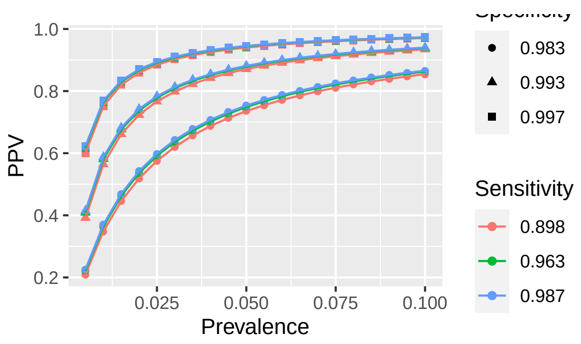


**(a)** Luminex

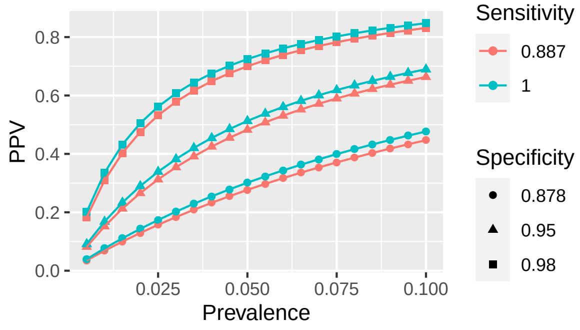


**(b)** Megna

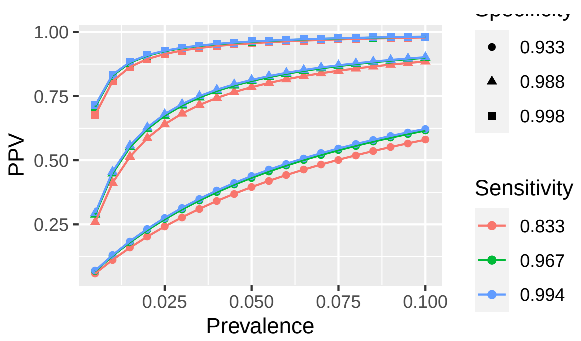


**(c)** NanoEnTek

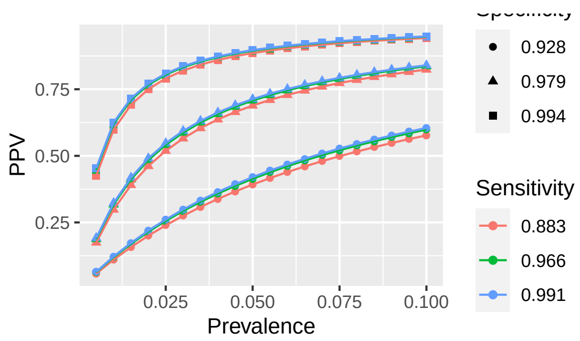


**(d)** Nirmidas

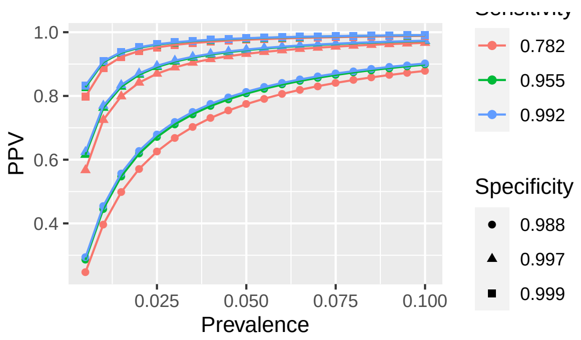


**(e)** QuanSys

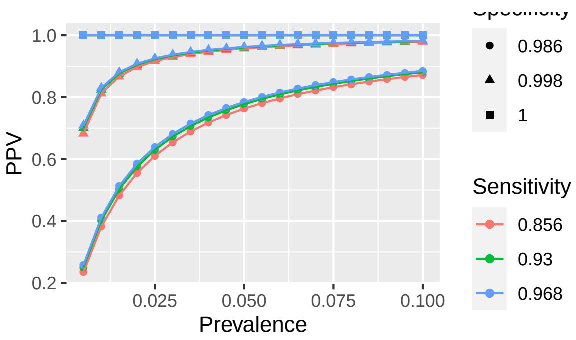


**(f)** Quotient

**Figure S7.** EUA Tests: L-R

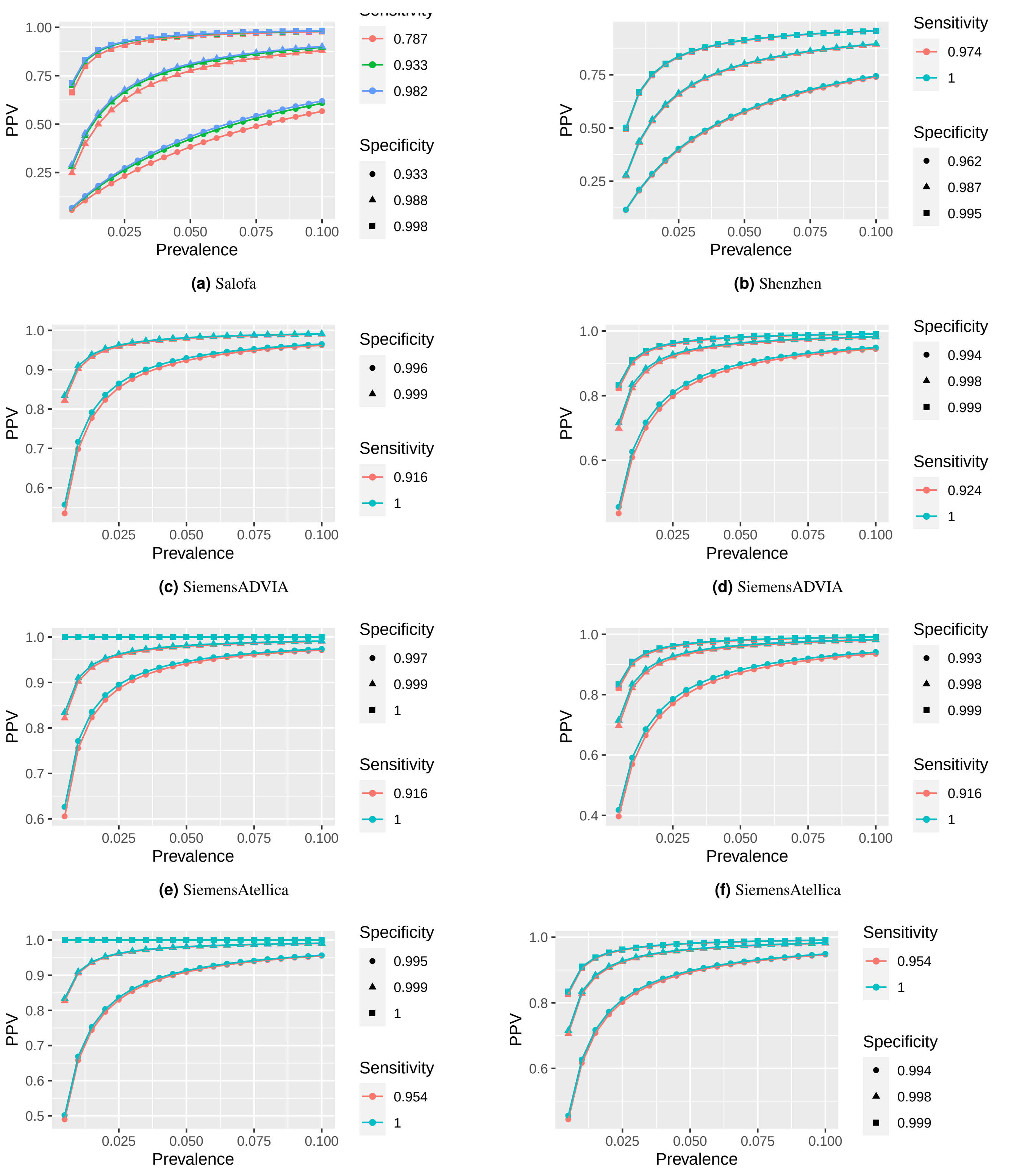


**(a)** Salofa **(b)** Shenzhen

**(c)** SiemensADVIA **(d)** SiemensADVIA

**(e)** SiemensAtellica **(f)** SiemensAtellica

**(g)** SiemensDimEXL **(h)** SiemensDimVista

**Figure S8.** EUA Tests: Sa-Si

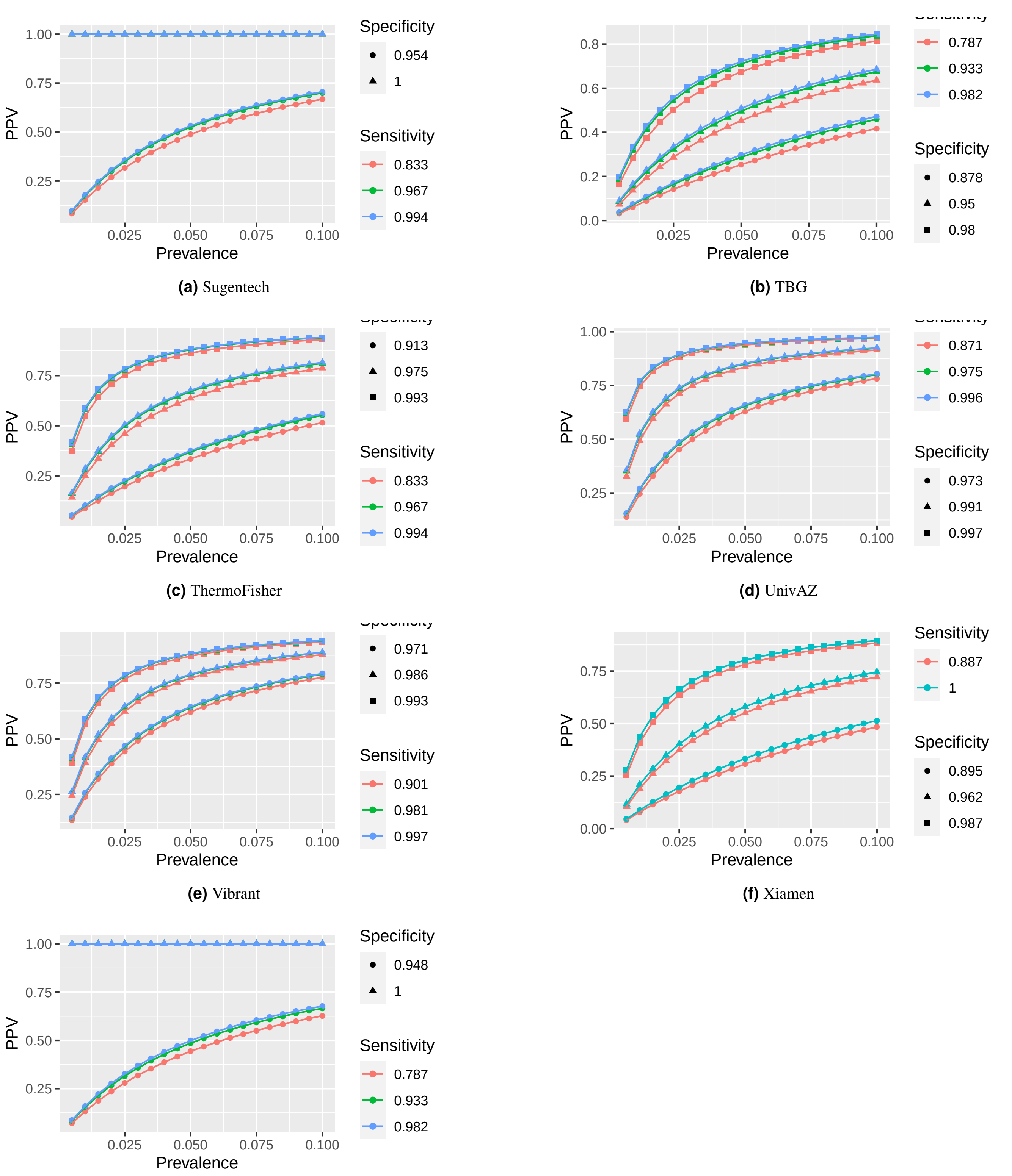


**(a)** Sugentech

**(b)** TBG

**(c)** ThermoFisher

**(d)** UnivAZ

**(e)** Vibrant

**(f)** Xiamen

**(g)** Zeus

**Figure S9.** EUA Tests: Su-Z
